# Trajectories of brain structure and function in young adult carriers of genetic frontotemporal dementia variants

**DOI:** 10.64898/2026.06.08.26355165

**Authors:** Isis So, Jolina Lombardi, Adam M. Staffaroni, Kristy K. L. Coleman, Arabella Bouzigues, Eve Ferry-Bolder, Eva Cullen, Lucy L. Russell, Phoebe H. Foster, Sophie Farley, Rhian Convery, John C. van Swieten, Lize C. Jiskoot, Harro Seelaar, Daniela Galimberti, Rik Vandenberghe, Robert Laforce, Rose Bruffaerts, Maxime Bertoux, Thibaud Lebouvier, Eino Solje, Johannes Levin, Giuseppe di Fede, Alexander Thompson, Isabelle Le Ber, Raffaella Lara Migliaccio, Peter Körtvélyessy, Matthias L. Schroeter, Giancarlo Logroscino, Markus Otto, Zeljko Uzelac, Ignacio Illán-Gala, Johanna Krüger, Benedetta Nacmias, Alexander Gerhard, Tobias Langheinrich, Simon Ducharme, Isabel Santana, Maria Carmela Tartaglia, Mario Masellis, Alexandre de Mendonça, James B. Rowe, Caroline Graff, Fermin Moreno, Matthis Synofzik, Barbara Borroni, Raquel Sanchez-Valle, Danielle Brushaber, Ciaran M. Considine, Kelley Faber, Anne Fagan, Julie A. Fields, Tatiana Foroud, Leah K. Forsberg, Tania Gendron, Daniel Geschwind, Hilary W. Heuer, Eric Huang, Kejal Kantarci, Tyler Kolander, Argentina Lario Lago, Shannon B. Lavigne, Carly Mester, Joie Molden, Leonard Petrucelli, Rosa Rademakers, Eliana Marisa Ramos, Katherine P. Rankin, Katya Rascovsky, Kristoffer W. Rhoads, Marijne Vandeberghe, Sandra Weintraub, Bonnie Wong, Bradley F. Boeve, Adam L. Boxer, Howard J. Rosen, Suzee E. Lee, Jonathan D. Rohrer, Elizabeth C. Finger, the Frontotemporal Dementia Prevention Initiative (FPI) Investigators

## Abstract

**Background and Objectives:** Converging evidence hints at neurodevelopmental effects in genetic frontotemporal degeneration (FTD). In cross-sectional studies, for some genes, young adult FTD variant carriers show differences in brain volumes and cognition compared to familial non-carriers. However, longitudinal trajectories may more sensitively capture FTD-related neurodevelopmental vs. neurodegenerative changes than cross-sectional approaches. This study examined longitudinal trajectories of brain volumes, executive function, and plasma biomarkers in young adult carriers compared to familial non-carriers, as measures of neurodevelopmental and neurodegenerative outcomes of FTD-causing variants.

**Methods:** This longitudinal cohort study comprised participants, aged 18-30 years, from the FTD Prevention Initiative across Europe, Canada, and the USA. Genetic groups included *C9orf72* (47%), *MAPT* (30%), and *GRN* (23%). Linear mixed-effects models were computed to assess longitudinal outcomes across age between groups, controlling for sex, scanner (for brain volumes), and education (for executive function); random effects accounted for between-subject variability nested within family membership.

**Results:** Variant carriers (*n*=147) and familial non-carriers (*n*=113) did not differ in age (mean±SD, 25.9±3.2 years), sex (53% female), or number of visits (2.1±1.7). Young adult *C9orf72* repeat expansion carriers exhibited smaller thalamic volumes than non-carriers at the reference age of 26 years (*b*=-982.8mm^3^, SE=317.0, *p=*0.0046, *f*^2^=0.32), with relatively stable trajectories across ages 18-30 (i.e., no change over time). Trajectories of rostral anterior cingulate volumes differed in *C9orf72* carriers and non-carriers across age, where carriers showed relatively stable trajectories and non-carriers showed age-appropriate declines (*b*=64.4mm^3^, SE=29.9, *p=*0.035, *f*^2^=0.07). For *MAPT* and *GRN*, there were little to no differences in total brain, cortical, or subcortical volumes between groups and over time. No longitudinal differences were observed between carriers and non-carriers in executive function, or plasma NfL or GFAP for any genetic group.

**Discussion:** *C9orf72* repeat expansions were linked to smaller average thalamic volumes and stable trajectories between ages 18 to 30, supporting potential neurodevelopmental origins. The modest evidence supporting an absence of difference in neurodegenerative biomarkers and executive function suggests minimal early neurodegeneration and functional preservation in young adulthood.

## INTRODUCTION

Frontotemporal degeneration (FTD) is widely recognized as a dementia that starts in mid-life or later.^1^ However, converging preclinical^2–11^ and human neuroimaging^12–17^ studies have identified group-level differences decades before expected symptom onset, raising the possibility of neurodevelopmental effects in genetic forms of FTD. The most common genes in which variants cause genetic FTD are *C9orf72, GRN*, and *MAPT*.^1^ These genes are expressed and active during pre- and post-natal neurodevelopmental stages,^18^ contributing to neural stem cell and progenitor proliferation, synaptic connectivity, plasticity, and pruning, and cortical neuron maturation.^2–11^ Slight differences in total intracranial volume (a neurodevelopment marker stable in adult ages) have been observed in *GRN* and *MAPT* carriers,^16^ as has reduced gyrification (a neurodevelopmental process which begins during first trimester and peaks during childhood) in asymptomatic *C9orf72* carriers.^14^ The field has also increasingly raised the idea of characterizing genetic FTD across the lifespan to better understand mechanisms of resiliency and vulnerability to disease.^19^

Typical neurodevelopment across youth and young adulthood is usually modelled longitudinally,^20–22^ but neurodevelopment can have high individual heterogeneity, which presents a challenge to interpreting group-level analyses. An attempt to overcome this limitation resulted in the development of age- and sex-normed models of brain growth, specific to subcortical volumes, cortical thickness, and surface area.^23^ Conceptually analogous to pediatric growth charts, normative models provide a reference against which individual brain morphometry metrics can be compared.^24^ Normative models demonstrate that typical neurodevelopment between 18-30 years old is associated with mild linear declines in subcortical volumes and cortical thickness in regions commonly affected in FTD.^23^

International cohort studies have enabled characterization and predictive modelling of neurodegeneration in genetic FTD across presymptomatic to symptomatic stages, primarily between 40-80 years old. During presymptomatic stages, the regions which first demonstrate volume differences include the thalamus in *C9orf72*-associated FTD,^12,19,25^ rostral anterior cingulate in *GRN*-associated FTD, ^12,19,25^ and the temporal pole, hippocampus, and amygdala in *MAPT*-associated FTD.^19,25,26^ Disease progression models suggest that executive function differences may occur 10-40 years before symptom onset in *C9orf72* and *GRN* carriers, and up to 10 years in *MAPT* carriers.^25^ However, trajectories of behavioural and clinical symptoms do not differ in presymptomatic variant carriers for all genetic groups compared to non-carriers.^27^ Plasma neurofilament light chain (NfL), a marker of axonal injury and neurodegeneration, has been modelled to deviate from normal levels as early as 30 years before disease onset in *C9orf72* carriers, and up to 15 years in *GRN* carriers,^25^ but has not been examined in young adults. These studies collectively highlight the question of whether early differences represent altered neurodevelopment or early neurodegeneration. Longitudinal investigations in young adult FTD variant carriers are lacking, but are key for sensitively capturing FTD-related changes that may be masked in cross-sectional analyses, and for differentiating whether changes are more likely attributable to neurodevelopmental or early neurodegenerative processes.

This study examined longitudinal trajectories of brain volumes, executive function, behavioural measures, and plasma biomarkers in young adult carriers compared to familial non-carriers, as measures of neurodevelopmental and neurodegenerative outcomes of FTD-causing variants. Compared to their respective young adult non-carriers, FTD variant carriers were expected to show longitudinal brain volume differences, but no change in cognitive or behavioural function due to potential ability to compensate during younger ages,^17^ nor differences in neurodegenerative biomarkers. As an exploratory objective, to better account for inter-individual heterogeneity during neurodevelopment, normative models were used to derive z-scores of individual variations in brain morphometry metrics to further compare young adult carrier and non-carrier participants.

## METHODS

### Participants

This study included 260 young adults enrolled in the Genetic Frontotemporal Dementia Initiative (GENFI:https://www.genfi.org/), or ARTFL-LEFFTDS Longitudinal Frontotemporal Lobar Degeneration Study (ALLFTD: https://www.allftd.org/, NCT04363684; ARTFL=Advancing Research and Treatment for Frontotemporal Lobar Degeneration study: NCT02365922; LEFFTDS=Longitudinal Frontotemporal Lobar Degeneration study, NCT02372773). GENFI phases 1-3 (GENFI1, 2012-15; GENFI2, 2015-21; GENFI3, 2021-26) included 46 clinical research sites across Europe and Canada. ALLFTD included 28 sites across the USA and Canada. Data comprised GENFI Data Freeze 8 and the ALLFTD Data Freeze dated August 22, 2025.

The full inclusion and exclusion criteria for GENFI and ALLFTD have been outlined elsewhere.^25,26^ Participants were carriers or familial non-carriers of known pathogenic genetic variants in *C9orf72* (>30 repeats), *GRN*, or *MAPT*; mutation lists for *GRN* and *MAPT* are listed on the GENFI website. A specific inclusion criterion for this study included young adults, aged 18-30 years. Ethics approval was obtained per individual site. Written informed consent was provided by all participants, in line with the Declaration of Helsinki.

### Study Design and Procedures

GENFI and ALLFTD are prospective, longitudinal, and observational studies, which gather demographics, neuroimaging, neuropsychological, behavioural, and clinical outcomes typically at 1-to-2-year intervals. This study examined brain volumes and cortical thickness from T1-weighted MRI, executive function task performance, behavioural measures, and plasma GFAP and NfL in genetic variant carriers in comparison to familial non-variant carriers.

### Neuroimaging

Details of neuroimaging protocols for GENFI and ALLFTD are found in prior reports.^25,26^ GENFI participants completed T1-weighted MRI on a 1.5T (Siemens, GE) or 3T scanner (Siemens Trio, Siemens Skyra, Siemens Prisma, Philips Achieva, GE Discovery MR750). Sequence parameters included: 256 x 256 x 208 matrix; 208 slices; 1.1 mm isotropic voxel size; 8° flip angle; TR and TE varied by vendor. ALLFTD participants completed T1-weighted MRI on 3T scanners (GE, Philips, Siemens). Magnetization Prepared Rapid Gradient Echo (MPRAGE) image parameters included: 240 x 256 x 256 matrix; 170 slices; voxel size=1.05 x 1.05 x 1.25 mm^3^; flip angle, TR, and TE varied by vendor.

All images were visually inspected for quality control by the respective core imaging groups in GENFI and ALLFTD, and scans with excessive movements or artifacts were removed. Preprocessing was conducted in FreeSurfer v7.4.1,^28^ and the Desikan-Killiany^29^ and Aseg^30^ atlases were used for cortical and subcortical volumetric segmentation, respectively.

Regions of interest (ROI) for all genetic groups included volumes of the total brain, medial orbitofrontal cortex (mOFC), and frontal pole. Additional bilateral ROI differed by genetic group and were selected based on established regions of early involvement in older pre-symptomatic and symptomatic adults.^25,26^ This included the: thalamus, insula, and rostral anterior cingulate cortex (rACC) for *C9orf72*; insula, rACC, and putamen for *GRN*; and anterior medial temporal cortex (encompassing the hippocampus, amygdala, parahippocampal and entorhinal cortices, and temporal pole) for *MAPT*.

### Executive Function Measures

Executive function tests that were common between the GENFI Neuropsychology Battery^26^ and the National Alzheimer’s Coordinating Center Uniform Data Set v3.0 Neuropsychology Battery used in ALLFTD^31^ were used to calculate a composite score. This included Verbal Fluency (Animals), Verbal Fluency (Letter F), Digit Span Backward (maximum number of consecutive digits correctively produced), and Trail Making Tests-A and B. The composite calculation followed Staffaroni et al. (2021),^32^ with two modifications made for the present study. First, as Verbal Fluency (Vegetables) and Verbal Fluency (Letter L) were not available from most participants, factor loading was adjusted accordingly to avoid inclusion of these tests. Second, rather than directly accounting for age, sex, and education effects per the original composite (which was validated in participants aged 60-70s with non-linear age normalization), these demographic variables were included as covariates in our executive function models.

### Behavioural Measures

Behavioural outcomes were assessed using informant-based questionnaires: the Cambridge Behavioural Inventory-Revised (CBI-R)^33^ in GENFI, and the Neuropsychiatric Inventory Questionnaire (NPI-Q)^34^ in ALLFTD.

The CBI-R evaluates the presence and frequency of the following behaviours: memory and orientation, everyday skills, self-care, abnormal behaviour, mood, beliefs, eating habits, sleep, stereotypical and motor behaviours, and motivation.^33^ Likert-scale scores range from 0 to 4, with higher scores representing greater frequency of a behaviour in the past month.^33^ The NPI-Q measures the presence (yes/no) and severity (mild/moderate/severe) of 12 behaviours: delusions, hallucinations, agitation or aggression, depression or dysphoria, anxiety, elation or euphoria, apathy or indifference, disinhibition, irritability or lability, motor disturbance (including repetitive activities), nighttime behaviours, and appetite and eating.^34^

The following behaviours were of interest to neurodevelopmental outcomes in FTD, and were measured by both the CBI-R and NPI-Q: disinhibition, apathy, empathy, obsessive-compulsive disorder, appetite changes, delusions, hallucinations, depression, anxiety, aggression.

### Plasma NfL and GFAP

Plasma collection, processing, and storage procedures have been reported for GENFI^35^ and ALLFTD.^25,36^ Plasma aliquots were stored at -80°C prior to analysis. Plasma NfL and GFAP concentrations were quantified according to manufacturer protocols at a respective core biomarker site per consortia, and analyses were performed by a single operator blinded to genetic and clinical information. In GENFI, plasma NfL and GFAP concentrations were quantified using the Simoa Neurology 4-Plex A assay (Quanterix, Cat. No. 102153) on the Quanterix HD-1 Analyzer; assay sensitivity was 0.221 pg/mL for GFAP and 0.104 pg/mL for NfL. In ALLFTD, plasma NfL concentrations were measured using the Simoa NF-Light digital immunoassay (Quanterix, Cat. No. 103186), and plasma GFAP using the Simoa GFAP digital immunoassay (Quanterix, Cat. No. 102336, Lot 503909), both on the HD-X Analyzer. In ALLFTD, NfL was assayed using two kit lots, with measurements derived from kits corresponding to Lot 501992 and Lot 503729, and additional samples measured across both lots to support inter-assay normalization. Across both consortia, assay performance was monitored using manufacturer-provided calibrators and quality control samples, with duplicate measurements and repeat analyses performed as needed to ensure reliability and consistency across runs.

### Statistical Analysis

Statistical analyses were computed with R v4.4.1. Demographic variables were summarized using means and standard deviations for continuous variables, or participant counts for categorical variables. Demographic comparisons between groups were conducted with t-tests or chi-square tests for continuous and categorical variables, respectively. Given the low frequency of FTD-related behaviours in young presymptomatic carriers,^12,27^ CBI-R and NPI-Q symptom endorsement was categorized as if ever present (present) or never present (absent), and group differences were examined using chi-square tests. Statistical assumptions for linear mixed-effects (LME) models included linearity, normally distributed residuals, and homogeneity of residual variance; these assumptions were evaluated using Q-Q plots and histograms and passed prior to model computation. Model multicollinearity was assessed using variance inflation factor tests and determined acceptable.

LME models, computed separately for *C9orf72, GRN*, and *MAPT*, compared longitudinal outcomes between carriers and non-carriers in brain volumes, executive function composite scores, and plasma biomarkers of neurodegeneration via NfL and GFAP levels. For all models, fixed effects included group, age, sex, and a group-by-age interaction; random effects accounted for between-subject variability nested within family membership. Age was centered at the sample mean to facilitate interpretation of group effects. Scanner and education differences were also included as fixed effects in the models for brain volumes and executive function, respectively. Outliers (±3 standard deviations within a participant’s carrier status group) per outcome variable were removed prior to analyses. Benjamini-Hochberg corrections were applied for multiple comparisons within each genetic group for the neuroimaging ROIs, with α=0.05. Effect size was computed using Cohen’s *f*^2^, where 0.02, 0.15, and 0.35 represent small, medium, and large effects, respectively.^37^

Sensitivity analyses were conducted for the brain volume and executive function models. For the brain volume analyses, separate LME models controlled for total intracranial volume and *TMEM106B-rs1990622* genotype (coded as 0, 1, or 2 copies of the G allele). For the executive function analyses, a separate model included visit site to account for potential cultural and educational differences that may affect cognitive testing. Further sensitivity analysis determined whether the main effects of interest were present specifically in the ALLFTD cohort, as some GENFI participants had been previously examined cross-sectionally,^12^ and with participants with only one visit removed.

For volumetric ROIs that showed a significant main effect of group or group-by-age interaction, post-hoc models were computed, with a model fitted separately for the left and right ROI to explore possible laterality effects. For cortical volume ROIs with significant effects, a post-hoc model was fitted for cortical thickness to assess whether the observed volume effects reflected underlying gray matter differences.

### Normative Modelling for Neuroimaging ROIs

As an exploratory analysis, we used normative models to quantify deviations from typical development in our young adult FTD cohort. Normative models of brain growth, developed by the ENIGMA Lifespan Working Group, characterized age- and sex-normed trajectories of brain morphometry across development and aging. These models were generated using MRI data from 37,407 healthy individuals aged 3 to 90 years (53.4% female) across 87 international research sites, and published as an open-source reference, CentileBrain (https://centilebrain.org/).^23^ Normative models were adjusted for age, sex, total intracranial volume, and scanner manufacturer. Regions were derived from FreeSurfer’s Aseg atlas for subcortical volume, and the Desikan-Killiany atlas for cortical thickness and surface area, with all regions modelled separately by hemisphere, resulting in 14 subcortical volume, 68 cortical thickness, and 68 surface area models.

Individual-level deviation scores (z-scores) were computed for all participants using CentileBrain for the same subcortical ROIs used in the primary analysis. When a normative model was unavailable for a given subcortical ROI, cortical thickness of the anatomically corresponding cortical region was used instead. All ROIs in the normative-modelling analyses were evaluated separately by hemisphere, per available normative models. Across all genetic groups, ROIs included cortical thickness of the mOFC and frontal pole. Additional ROI, per our primary analyses, differed by genetic group based on established regions affected in symptomatic FTD: *C9orf72* included thalamic volumes, and cortical thickness of the insula and rACC; *GRN* included putamen volumes, and cortical thickness of the insula and rACC; and *MAPT* included hippocampal and amygdala volumes, and cortical thickness of the frontal pole, mOFC, temporal pole. LME models were then used to evaluate longitudinal differences between young adult FTD variant carriers and non-carriers, with normative deviation z-scores as the outcome, and a participant-level random intercept to account for repeated measures.

### Data Availability

Data requests can be made by an FPI-affiliated investigator to the coordinating centre. Preprocessing and analytical scripts can be shared upon request.

## RESULTS

### Participant Demographics

On average, participants (*n*=260) ranged from 18-30 (mean±SD: 25.9±3.2) years old, attended 2.1±1.7 visits, and had 14.9±2.3 years of education **(Table 1)**. Demographic variables (age, number of visits, sex, years of education, and handedness) did not differ between young adult carriers and non-carriers for *C9orf72, GRN, MAPT* (all p>0.05), except in *C9orf72* time between visits (*p*=0.004, *d*=-0.66) and frequency of handedness (*p*=0.003, *V*=0.23).

**Table 1.** Demographic characteristics of all participants.

| Characteristic | All |  |  | *C9orf72* |  |  | *GRN* |  |  | *MAPT* |  |  |
| --- | --- | --- | --- | --- | --- | --- | --- | --- | --- | --- | --- | --- |
|  | Total | Carriers | Non-carriers | Carriers | Non-carriers | Between group comparison | Carriers | Non-carriers | Between group comparison | Carriers | Non-carriers | Between group comparison |
| Sample size (*n*) | 260 | 147 | 113 | 72 | 48 | NA | 35 | 26 | NA | 40 | 39 | NA |
| Number of visits (mean ± SD) | 2.1 ± 1.7 | 2.2 ± 1.7 | 2.1 ± 1.5 | 1.9 ± 1.6 | 1.7 ± 0.9 | $t=1.12$,<br>$p=0.27$,<br>$d=0.19$ | 1.9 ± 1.3 | 2.3 ± 2.0 | $t=-0.96$,<br>$p=0.34$,<br>$d=-0.26$ | 3.0 ± 2.1 | 2.4 ± 1.8 | $t=1.18$,<br>$p=0.24$,<br>$d=0.27$ |
| Time between visits, years (mean ± SD) | 1.37 ± 0.63 | 1.34 ± 0.60 | 1.41 ± 0.68 | 1.28 ± 0.43 | 1.77 ± 1.06 | $t=-3.04$,<br>$p=0.004^{**}$,<br>$d=-0.66$ | 1.30 ± 0.48 | 1.48 ± 0.48 | $t=-1.45$,<br>$p=0.15$,<br>$d=-0.38$ | 1.41 ± 0.75 | 1.16 ± 0.34 | $t=1.92$,<br>$p=0.06$,<br>$d=0.43$ |
| Age, years (mean ± SD) | 25.9 ± 3.2 | 25.9 ± 3.3 | 26.0 ± 3.1 | 26.4 ± 3.1 | 27.0 ± 2.5 | $t=-1.59$,<br>$p=0.11$,<br>$d=-0.22$ | 26.3 ± 2.8 | 26.3 ± 2.6 | $t=-0.08$,<br>$p=0.94$,<br>$d=-0.01$ | 25.1 ± 3.5 | 24.9 ± 3.5 | $t=0.30$,<br>$p=0.77$,<br>$d=0.04$ |
| Ratio of participants with 2+ visits / total (*n*):<br>Neuroimaging<br>Executive function<br>Plasma biomarkers | 135/260<br>133/259<br>71/118 | 77/147<br>76/146<br>38/59 | 58/113<br>57/113<br>33/59 | 32/72<br>33/71<br>11/25 | 22/48<br>22/48<br>9/21 | NA | 16/35<br>14/35<br>8/16 | 13/26<br>12/26<br>8/16 | NA | 29/40<br>29/40<br>19/28 | 23/39<br>23/39<br>16/22 | NA |
| Sex (*n*)<br>female: male | 142: 118 | 82: 65 | 60: 53 | 40: 32 | 28: 20 | $\chi^2=0.002$,<br>$p=0.96$,<br>$V=0.003$ | 19: 16 | 13: 13 | $\chi^2=0$,<br>$p=1$,<br>$V=0$ | 23: 17 | 19: 20 | $\chi^2=0.24$,<br>$p=0.62$,<br>$V=0.03$ |
| Education, years (mean ± SD) | 14.9 ± 2.3 | 15.1 ± 2.2 | 14.7 ± 2.4 | 15.3 ± 2.1 | 14.9 ± 2.7 | $t=0.64$,<br>$p=0.52$,<br>$d=0.09$ | 15.1 ± 1.9 | 14.8 ± 2.5 | $t=0.63$,<br>$p=0.53$,<br>$d=0.11$ | 14.7 ± 2.0 | 14.3 ± 2.3 | $t=0.52$,<br>$p=0.61$,<br>$d=0.07$ |
| Handedness (*n*)<br>Right<br>Left<br>Ambidextrous | >200<br>10-50<br><10 | 100-150<br>10-50<br><10 | 50-100<br>10-50<br><10 | 50-100<br><10<br><10 | 10-50<br><10<br><10 | $\chi^2=11.9$,<br>$p=0.003^{**}$,<br>$V=0.23$ | 10-50<br><10<br><10 | 10-50<br><10<br><10 | $\chi^2=0.70$,<br>$p=0.40$,<br>$V=0.07$ | 10-50<br><10<br><10 | 10-50<br><10<br><10 | $\chi^2=2.57$,<br>$p=0.11$,<br>$V=0.11$ |
| Sites (*n*) | 38 | 35 | 31 | 27 | 26 | NA | 19 | 11 | NA | 16 | 14 | NA |

**\*\*** $p<0.01$ between carriers and non-carriers. Handedness counts are reported in ranges to prevent unblinding participants who may be left-handed or ambidextrous.
Abbreviations: SD, standard deviation; NA, not applicable.

### Neuroimaging

#### C9orf72

Brain volume trajectories of six ROIs were examined between young adult *C9orf72* carriers and non-carriers, with significant trajectory differences observed in the bilateral thalamus and rACC, and trends towards significance in the mOFC and total brain **(Figure 1)**. Models of the frontal pole and insula for *C9orf72* yielded no group effects or group-by-age interactions. Specifically, young adult *C9orf72* carriers exhibited smaller thalamic volumes than non-carriers (*b*=-982.8mm^3^, SE=317.0, *p=*0.0046, *f*^2^=0.32), with large effect size and surviving Benjamini-Hochberg correction. No significant group-by-age interaction was observed in the thalamus (*p*=0.98).

**Figure 1.**
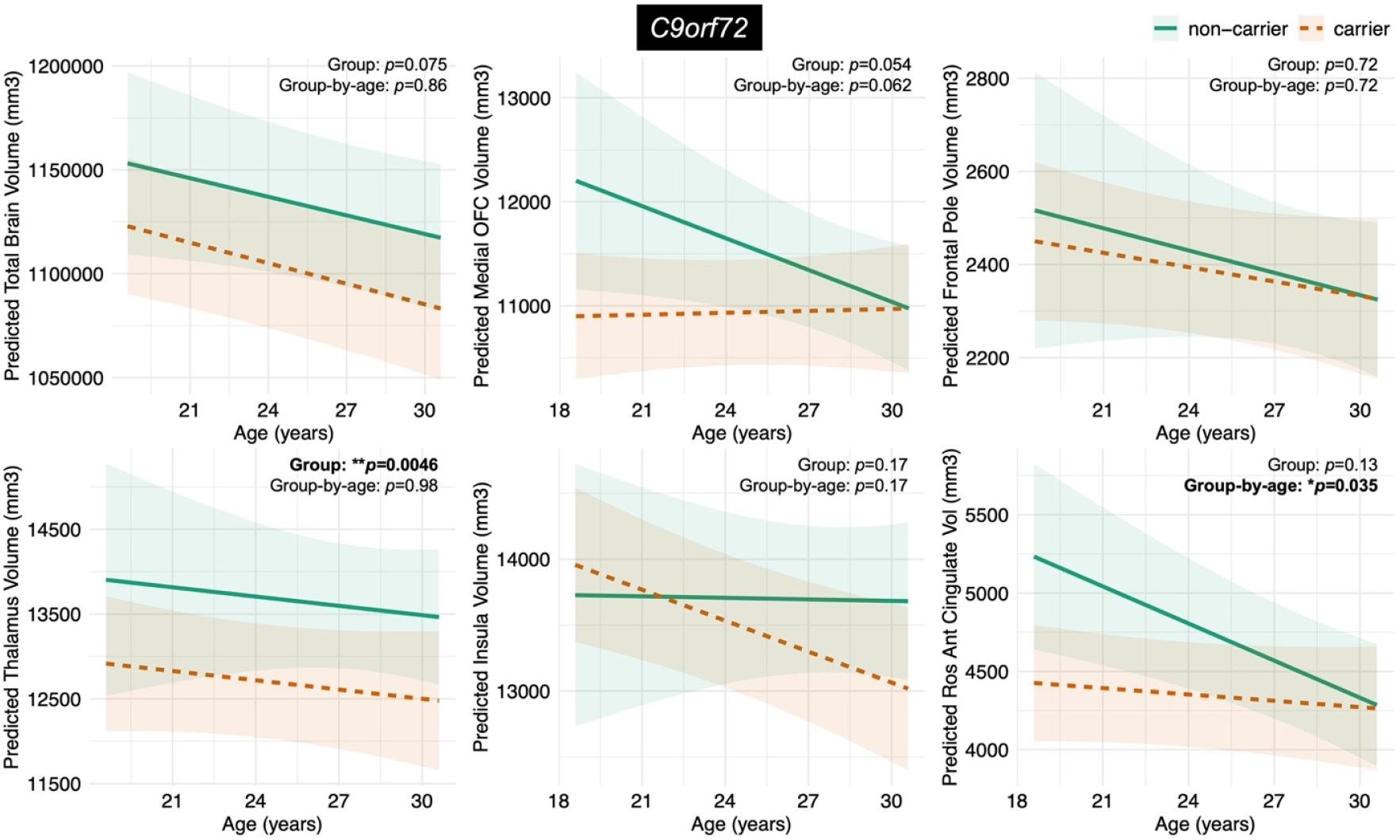
Predicted trajectories of young adult *C9orf72* repeat expansion carriers compared to non-carriers for all regions of interest. On average, young adult *C9orf72* repeat expansion carriers demonstrated smaller thalamic volumes and trended towards smaller total brain volumes compared to non-carriers. Young adult carriers also showed relatively stable trajectories in the rACC and mOFC across ages 18 to 30 compared to non-carriers, who exhibited expected age-related declines. No main effects of group or differences in longitudinal trajectories were observed in the frontal pole and insula. The plotted confidence intervals reflect pointwise uncertainty in the predicted marginal means for each group and do not represent uncertainty in the group-by-age interaction term, which tests differences in age-related slopes. Abbreviations: rACC = rostral anterior cingulate cortex; mOFC = medial orbital frontal cortex.

There was no difference in rACC volumes between carriers and non-carriers at the sample reference age (*b*=-288.7mm^3^, SE=186.58, *p*=0.13, *f*^2^=0.06). However, a significant group-by-age interaction in the rACC (*b*=64.4mm^3^, SE=29.9, *p=*0.035, *f*^2^=0.07) revealed that young adult *C9orf72* non-carriers showed age-related volume reductions (*b*=-78.1mm^3^/year, 95% CI [-127.9, -28.3]), whereas carriers showed relatively stable trajectories over time (+13.7mm^3^/year, 95% CI [-44.8, 17.4]). A post-hoc model which fitted bilateral rACC cortical thickness as the outcome variable found no significant group-by-age interaction or main effect of group (*p*>0.05). Other post-hoc models, which explored left and right thalamic and rACC volumes separately, revealed consistent patterns **(Supplementary Material)**.

#### GRN

No main effects of group, nor group-by-age interactions, were observed in the total brain, frontal pole, mOFC, insula, rACC, or putamen **(eTable 2)**.

#### MAPT

For total brain volume, a group-by-age interaction was observed with small effect size (*b*=-2882.2mm^3^, SE=1222.4, *p*=0.02, *f*^2^=0.06), where young adult *MAPT* carriers demonstrated faster rates of decline (*b*=-3332.0mm^3^, 95% CI [-4996, -1667]) compared to non-carriers (*b*=-282.0mm^3^, 95% CI [-2607, 2044]) **(eTable 3)**. There were no differences in total brain and mOFC volumes between young adult *MAPT* carriers and non-carriers at the reference age (*p*>0.05 for both). Models of the frontal pole and anterior medial temporal cortex yielded no significant group-by-age interactions, nor main effects of group.

### Sensitivity Analyses

For all sensitivity analysis, including *TMEM106B-rs1990622* genotype, total intracranial volume adjustment, ALLFTD cohort only, and exclusion of single-visit participants, results were consistent with the primary analysis **(Supplementary Material)**.

### Normative Modelling Analysis with Neuroimaging ROIs

Normative deviation z-scores enabled greater control of inter-individual differences and provided additional insight into cortical and subcortical developmental trajectories per ROI, separated by hemisphere and sex. LME model contrasts for all genetic groups are reported in **eTable 4**. Significant results from *C9orf72* are described below.

Young adult male *C9orf72* carriers yielded smaller right thalamic volumes than non-carriers (*b*=-7.53, SE=2.91, *p*=0.013, *f*^2^ =0.07). The slopes of right thalamic volume trajectories differed (*b*=0.25, SE=0.11, *p*=0.026, *f*^2^ =0.25) in *C9orf72* young adult male carriers (*b*=0.15, 95% CI [0.035, 0.2675]) compared to non-carriers (*b*=-0.10, 95% CI [-0.29, 0.09]) **(Figure 2)**. In young adult female *C9orf72* carriers, right thalamic volumes in comparison to non-carriers trended but did not reach significance (*b*=4.52, SE=2.53, *p*=0.079, *f*^2^=0.05). Young adult male *C9orf72* carriers also showed thinner left mOFC in comparison to non-carriers (*b*=-5.08, SE=2.43, *p*=0.043, *f*^2^ =0.008) without slope differences.

**Figure 2.**
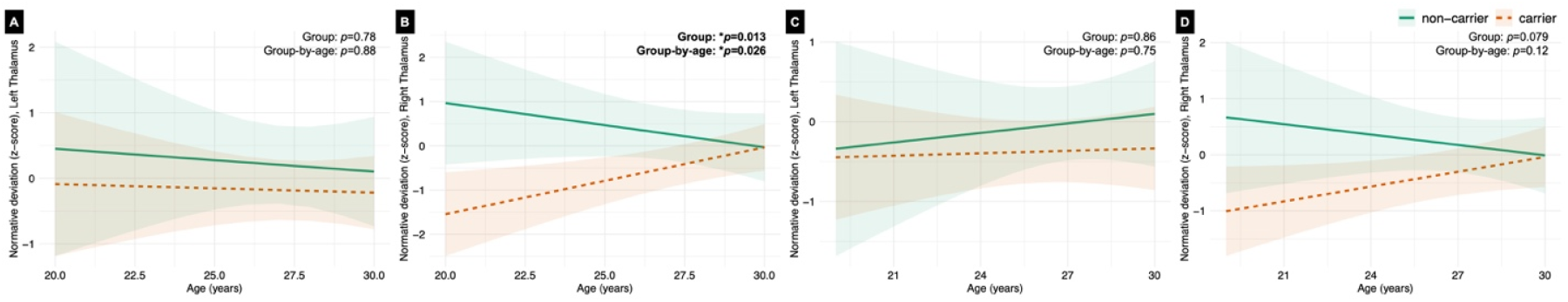
Predicted trajectories across age 18 to 30 per normative deviation z-scores of the thalamus between young adult *C9orf72* repeat expansion carriers and non-carriers. Models were computed separately for (**a**) left thalamus in males, (**b**) right thalamus in males, (**c**) left thalamus in females, and (**d**) right thalamus in females. Young adult male *C9orf72* expansion carriers had smaller thalamic volumes in the right thalamus at the mean sample age, and different right thalamic volume trajectories compared to non-carriers.

### Executive Function

Across all three genetic groups, there were no longitudinal differences in executive function composite scores of young adult variant carriers compared to non-carriers (group-by-age interactions all *p*>0.05, respectively) **(Figure 3)**. Per Bayes Factor analysis,^38^ the data for all gene groups were ∼3 times more likely reflective of the null hypothesis than the alternative **(eTable 5)**. Greater years of education was associated with greater executive function composite scores in all models (*C9orf72*: *p*=0.0001, *f*^*2*^ =0.18; *GRN*: *p*=0.012, *f*^*2*^ =0.11; *MAPT*: *p*=0.009, *f*^*2*^ =0.09). Sensitivity analyses, with site included as an additional covariate, yielded consistent findings.

**Figure 3.**
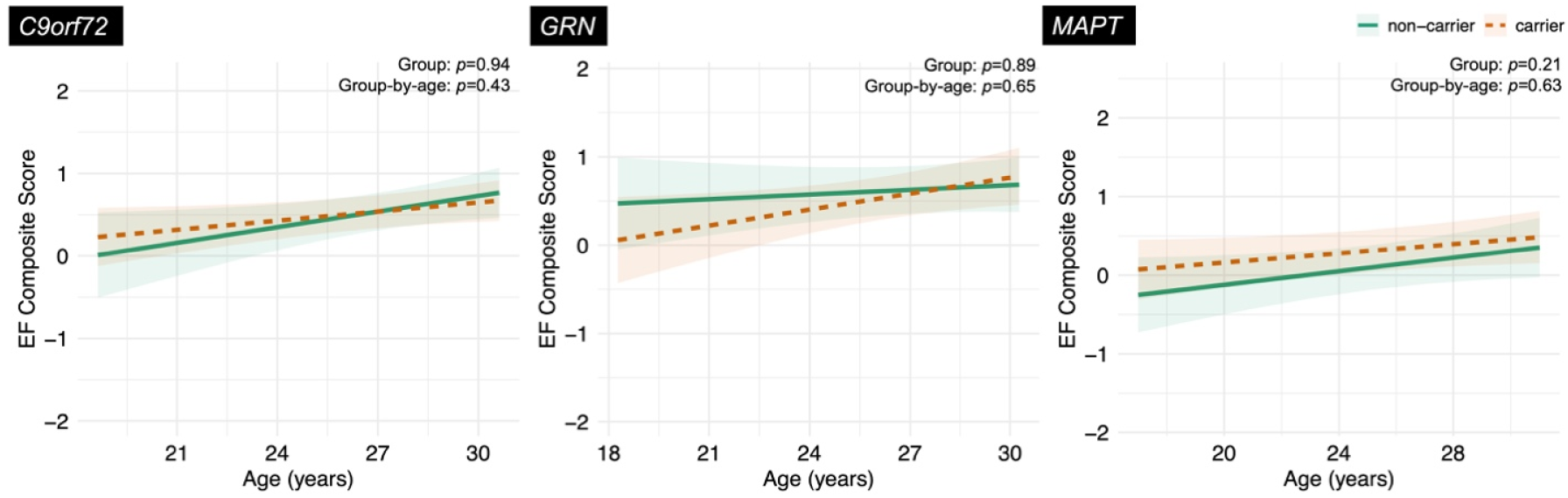
Predicted trajectories of young adult FTD variant carriers compared to non-carriers for executive function composite scores. No differences were observed in executive function composite scores across ages 18 to 30 between variant carriers and non-carriers of *C9orf72, GRN*, or *MAPT*.

### Behavioural Symptoms

Across all three genetic groups, young adult FTD variant carriers did not differ from non-carriers in the presence of disinhibition, apathy, empathy, obsessive-compulsive disorder, appetite changes, delusions, hallucinations, depression, anxiety, and aggression (chi-square statistics in **eTable 6**).

### Plasma NfL and GFAP

Across all three genetic groups, there were no significant group-by-age interactions or main effects for models of plasma GFAP and NfL (**Figure 4; eTable 5**).

**Figure 4.**
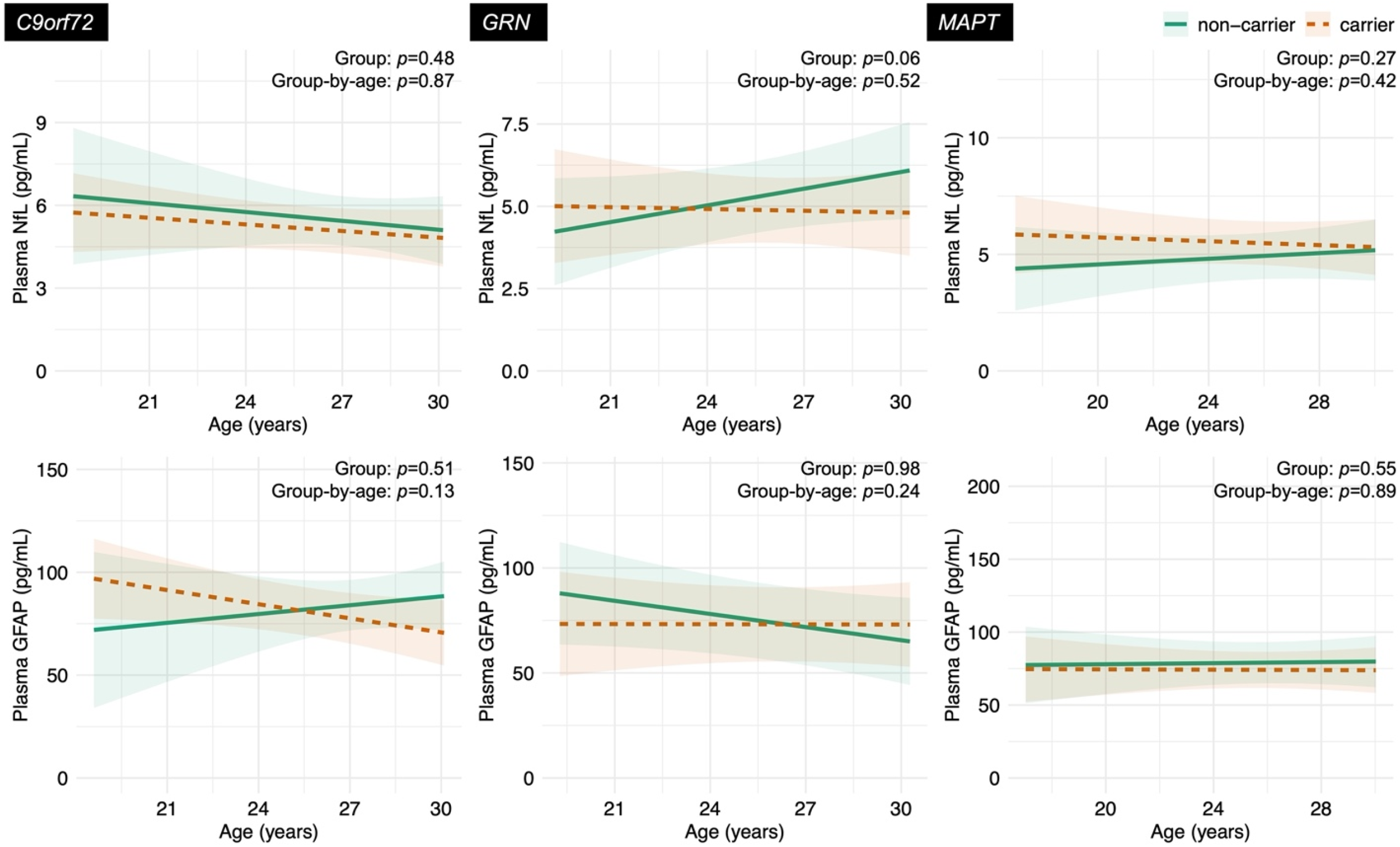
Predicted trajectories of young adult FTD variant carriers compared to non-carriers for plasma NfL (top panel) and GFAP (bottom panel). No differences were observed in plasma NfL or GFAP across ages *18 to* 30 between variant carriers and non-carriers of *C9orf72, GRN, or MAPT*.

## DISCUSSION

Longitudinal modelling of brain volumes between ages 18-30 suggests gene and region-specific findings for young adult FTD variant carriers compared to familial non-carriers. The lack of normative models available for cortical volumes prevented comparison of the same brain morphometry metric between our primary and exploratory analyses, but where available, yielded findings consistent with our primary results. This included the robust findings of smaller thalamic volumes in young adult *C9orf72* carriers, and trajectories which remained stable across 18-30 years old (no change over time) compared to expected first-order linear declines observed in non-carriers. In *C9orf72*-associated FTD, the earliest average age at which smaller thalamic volumes has been observed is 26 years,^12^ raising the question of whether such changes are more attributable to late neurodevelopmental or early neurodegenerative processes. Our *C9orf72* longitudinal models, which showed relatively stable volumetric slopes in carriers compared to non-carriers across all ROIs, coupled with a lack of between-group differences in NfL and GFAP, support a lack of neurodegeneration during young adulthood. Instead, smaller thalamic volumes in *C9orf72* carriers relative to non-carriers suggest that that these differences likely occurred before age 18 and persist during young adulthood.

These findings were reproduced by our normative model-derived deviation score analysis of thalamic volumes in *C9orf72* participants, which provided more stringent control of inter-individual variability. The sex-normed nature of the normative models provided additional insight into regional developmental trajectories separated by sex, though sex notably did not drive any differences in the primary analyses. While it is possible that the thalamic volume differences were driven by non-carriers having larger thalamic volumes than average neurodevelopment, all non-carrier volumes were within developmental norms and demonstrated expected mild linear age-related declines.^22,23^ Furthermore, relative to non-carriers, *C9orf72* carriers showed stable rACC trajectories across 18-30 years old, with smaller predicted rACC volumes preceding the reference age of 26, and a similar pattern trended towards significance in the mOFC. Similar to the significant thalamic volume findings, total brain volume followed a comparable trajectory in *C9orf72* carriers across young adulthood, but only at a trend level. Together, these *C9orf72* volume trajectory patterns raise the possibility that differences in some regions emerge before age 18, though this inference is indirect and not tested within the present cohort. The only ROIs where young adult *C9orf72* carriers and non-carriers were predicted to have similar volumes were the frontal pole and insula, consistent with models predicting later involvement of these regions.^19,25^

The model predictions of smaller volumes in *C9orf72* young adults align with preclinical data, where structural brain development, including thalamic volumes, has been shown to be vulnerable to *C9orf72* repeat expansions.^2,3^ For instance, *C9orf72* FTD/amyotrophic lateral sclerosis (ALS) patient-derived cerebral organoids were smaller in size and contained fewer CTIP2+ deep cortical layer neurons compared to healthy control-derived organoids.^2^ Embryonic transgenic mice with *C9orf72* repeat expansions also exhibited smaller thalamic and total brain volumes and reduced cortical thickness compared to wildtype mice, and *in vitro* human induced pluripotent stem cell models of *C9orf72*-associated FTD/ALS had reduced neural stem cell proliferation.^3^ Notably, typical human thalamic development begins during the first trimester of pregnancy, is orchestrated by complex spatiotemporal molecular patterns, and continues into young adulthood^39,40^; coupled with the fact that *C9orf72* is expressed and active during pre- and post-natal stages of human neurodevelopment,^18^ genetic alterations in *C9orf72* could affect a prominent time window of human neurodevelopment. Another question is whether *C9orf72* affects early cortical pruning, a process that extends into young adulthood. While no studies have examined the roles of C9orf72 on synaptic pruning during neurodevelopmental stages, loss of C9orf72 in motor cortex microglia can enhance synaptic pruning and neuroinflammation in knockout mouse models of FTD/ALS.^41^ Structural and functional MRI correlations in humans have posited functional network degeneration involving thalamic nuclei during both presymptomatic and symptomatic stages of *C9orf72*-associated FTD/ALS,^17,42^ though with compensatory effects which preserve cognition during presymptomatic stages.^17^ However, the field has yet to answer why the thalamus may be vulnerable to molecular and structural neurodevelopmental differences yet functionally resilient to FTD-related neurodegeneration until later life.

No differences in volume trajectories were detected in young adult *GRN* and *MAPT* variant carriers vs. non-carriers. Although young adult *MAPT* variant carriers appeared to show steeper declines in total brain volume than non-carriers across ages 18-30, this finding was confounded by wide confidence intervals and high within-group variances, and did not survive correction. Nevertheless, the *GRN* and *MAPT* longitudinal models may have been underpowered, limiting interpretability.

A potential confound to interpreting brain volume findings in FTD includes potential effects of the *TMEM106B-rs1990622* genotype, where the recessive G allele has been associated with larger cortical and subcortical volumes and better cognitive performance in presymptomatic *C9orf72* and *GRN* carriers, as well as reduced disease risk in presymptomatic *GRN* carriers.^43–45^ In our sensitivity analyses, controlling for *TMEM106B-rs1990622* yielded results consistent with subcortical and cortical volume trajectories in all genetic groups. However, there is little knowledge of whether *TMEM106B* affects neurodevelopment. A recent genome-wide association study conducted with data from over 11,600 American youths aged 9 to 17 flagged numerous genes associated with intracranial and subcortical volume development, but *TMEM106B* was notably not one.^46^ Only one study has directly examined the potential roles of *TMEM106B* in neurodevelopment, which revealed that the *TMEM106B* T186S variant in mice (comparable to the T185S variant in humans) promotes synaptic and axonal development in mouse primary hippocampal neurons at post-natal day zero.^47^ Overall, evidence for a role of *TMEM106B* in human neurodevelopment remains limited.

Trajectories of executive function composite scores and plasma NfL and GFAP levels yielded no significant differences between young adult FTD variant carriers and non-carriers, with Bayes Factor analyses providing some, albeit weak, evidence in support of the null hypothesis. Although prior cross-sectional findings in a smaller cohort observed increased executive function performance in young adult *GRN* and *MAPT* carriers (mean age: 26 years) compared to non-carriers,^12^ a common interpretation across those findings and current longitudinal ones is that variant carriers may be maintaining cognitive resiliency during young adulthood. Importantly, however, our longitudinal findings do not support that FTD genetic variants are associated with increased cognitive benefit in young adulthood. There were also no differences in the presence of FTD-related or clinical neurodevelopmental disorder-related behavioural symptoms across 18-30 years. This data is consistent with the null behavioural findings in FTD variant carriers in their mid-40s,^27^ and extends to younger adulthood, supporting that behavioural function is preserved until closer to disease onset. The earliest ages that plasma NfL and GFAP have been examined in FTD are the mid-40s,^35^ which is arguably outside the neurodevelopmental window that extends into the early 30s^22^; observations included elevated plasma NfL in presymptomatic *C9orf72* participants (mean age 44.2 years), and elevated plasma GFAP only in symptomatic *GRN* carriers (mean age 64.3 years), but no differences between NFL levels in presymptomatic *GRN* and *MAPT* carriers vs. non-carriers.^35^ Comparison of raw plasma NfL and GFAP values to normative reference ranges from a healthy Canadian cohort (*n*=900; age 3-79 years)^48^ suggests that our participants fall within normative ranges for young adulthood. Cumulatively, the modest evidence supporting a lack of difference in executive function, behaviour, and neurodegenerative plasma biomarkers, in the presence of brain volume differences in FTD variant carriers, suggest an absence of neurodegenerative change and persisting cognitive and behavioural resiliency during young adulthood.

Several limitations warrant consideration. Sample sizes were small in the *GRN* and *MAPT* groups for longitudinal modelling, though genetic FTD is a rare disease and young adult ages can be challenging to recruit. Cortical volumes were selected *a priori* as the primary structural metric to capture gray and/or white matter changes; however, cortical volume models could not be compared to a normative reference, which exist for cortical thickness but not volumes. Furthermore, the neuropsychological and behavioural measures used are validated and reliable in symptomatic adults with FTD, but may not be sensitive for young adults of otherwise typical development. CBI-R and NPI-Q subscales were employed as they enabled common variables between consortia, but are incomplete in reflecting symptoms and behaviours of clinical neurodevelopmental disorders. Future studies should use age-appropriate measures suitable for understanding atypical neurodevelopmental behaviours, including restricted interests, and early social, learning, or motor developmental differences, as has been deployed in GENFI-3.

Overall, this study observed differences between young adult *C9orf72* carriers and non-carriers in thalamic volume trajectories, including smaller thalamic volumes in carriers which remained stable across 18-30 years old. In conjunction with modest evidence of no differences in executive function composite scores, behaviour, and plasma NfL and GFAP levels between groups, these findings suggest that the observed volumetric differences more likely arise from neurodevelopmental rather than neurodegenerative processes. Given ongoing efforts to observe youth FTD variant carriers longitudinally, imminent studies should aim to understand the mechanisms that underlie functional resiliency in young adulthood. This could include correlations between structural and functional neuroimaging in young adult FTD cohorts, and preclinical models which examine how FTD variants may interact with cellular and molecular pathways of neurodevelopment. Understanding such pathways in human FTD may help identify new therapeutic targets that preserve functional independence or delay the onset of symptomatic disease.

## ACKNOWLEDGEMENTS

The authors thank all FPI cohort participants and caregivers for their efforts and contributions to this study. I.So and E.C.F. additionally thank Dr. Emma G. Duerden and Dr. J.Bruce Morton for their advice on modelling human neurodevelopment as it pertained to this study.

## APPENDIX Co-investigators

Co-investigators in the FPI are listed in the Supplementary Material.

