## Supplementary Material for "Trajectories of brain structure and function in young adult carriers of genetic frontotemporal dementia variants"

#### GENFI Investigators

Amelia Blesius,1 David Cash,1 Clementine Cheyne,1 Sophie Goldsmith,1 Luna Nordenstrom,1 Victoria Pauwels Romero,1 Hannah Rosenheim,1 David L. Thomas,2 Maura Malpetti,3 Alexander Murley,3 Sean Tan,3 Sonia Bellini,4 Valentina Cantoni,5 Roberto Gasparotti,6 Roberta Ghidoni,4 Enrico Premi,7 Andrea Arighi,8 Vittoria Borracci,8 Chiara Fenoglio,9 Manuela Pintus,8 Emanuela Rotondo,8 Luca Sacchi,8 Maria Serpente,8 Vittoria Aprea,10 Paola Caroppo,10 Marcella Catania,10 Giorgio Gelosa,10 Gemma Lombardi,10 Pietro Tiraboschi,10 Sandra Black,11 Morris Freedman,12 Sara Mitchell,11 Ekaterina Rogaeva,13 David Tang-Wai,14 Olaiya Aro,15 Liset de Boer,15 Julie De Houwer,15 Laura Donker Kaat,15 Elise Dopper,15 Jackie Poos,15 Babette Reichard,15 Tine Swartenbroekx,15 Valentina Bessi,16 Stefano Chiti,17 Giulia Giacomucci,16 Andrea Ginestroni,18 Valentina Moschini,18 Sonia Padiglioni,18 Andrew Douglas,19 Kevin Talbot,19 Martin Turner,19 Vesna Jelic,20 Elena Rodriguez-Vieitez,20 Melissa Taheri Rydell,20 Henrik Viklund,21 Caroline Dallaire-Théroux,22 Anna Antonell,23 Mircea Balasa,23 Sergi Borrego-Ecija,23 Paula Canasto,23 Jordi Junca,23 Albert Lladó,23 Tiago Costa-Coelho,24 Carolina Maruta,24 Gabriel Miltenberger,24 Ana Verdelho,24 Patricia Alves,25 Myriam Barandiaran,26 Ioana Croitoru,25 Alazne Gabilondo,26 Iliana Quiñones,25 Mikel Tainta,26 Lukas Beichert,27 Benjamin Bender,28 Lisa Graf,27 Mathieu Vandenbulcke,29 Koen Poesen,29 Sien Van Daele,29 Aline Delva,29 Maxime Montembeault,30 Guy Rouleau,31 Pierre-François Pradat,32 Giorgia Querin,33 Daisy Rinaldi,33 Anna Stockbauer,34 Alexander Bernhardt,34 Ellen Merli,34 Elisabeth Wlasich,34 Georg Nübling,34 Julia Kustermann,34 Olivia Wagemann,34 Sonja Schönecker,34 Lena Marth,34 Sarah Anderl-Straub,35 Ilse Dewachter,36 Stéphanie De Keulenaer,37 Rosa Rademakers,37 Marijne Vandebergh,37 Thijs Vande Vyvere,37 Sara Van Mossevelde,37 Xavier Delbeuck,38 Agnès Denève,38 Simon Lecerf,38 Juhana Hakumäki,39 Sanna Hannonen,39 Juho Kalapudas,39 Annemari Kilpeläinen,39 Ave Kivisild,40 Mervi Könönen,39 Inês Baldeiras,41 Miguel Castelo-Branco,41 João Durães,41 Marisa Lima,41 Maria Rosario Almeida,41 Miguel Tábuas-Pereira,41 Annerose Engel,42 Maryna Polyakova,42 Frank Regenbrecht,42 Angelika Thöne-Otto,42 Anna-Leena Heikkinen,43 Vesa Kiviniemi,43 Vesa Korhonen,43 Noora-Maria Suhonen,43 Laura Göschel,44 Jonah Nietiet,44 Anna Söllner,44 Teresa Dell'Abbate,45 Daniele Urso,45 Daniel Alcolea,46 Sara Bernal,46 Oriol Dols,46 Juan Fortea,46 Jesus García-Castro,46 Alberto Lleó,46 Sara Rubio-Guerra,46 Judit Selma-González,46

1. Department of Neurodegenerative Disease, Dementia Research Centre, UCL Queen Square Institute of Neurology, London, UK
2. Neuroimaging Analysis Centre, Department of Brain Repair and Rehabilitation, UCL Institute of Neurology, London, UK
3. Department of Clinical Neurosciences, University of Cambridge, Cambridge, UK
4. Molecular Markers Laboratory, IRCCS Istituto Centro San Giovanni di Dio Fatebenefratelli, Brescia, Italy
5. Department of Clinical and Experimental Sciences, University of Brescia, Brescia, Italy
6. Neuroradiology Unit, University of Brescia, Brescia, Italy
7. Stroke Unit, ASST Brescia Hospital, Brescia, Italy
8. Neurodegenerative Diseases Unit, Fondazione IRCCS Ca' Granda Ospedale Maggiore Policlinico, Milan, Italy
9. Department of Biomedical, Surgical and Dental Sciences, University of Milan; Fondazione IRCCS Ca' Granda Ospedale Maggiore Policlinico, Milan, Italy
10. Fondazione IRCCS Istituto Neurologico Carlo Besta, Milan, Italy

11. Sunnybrook Health Sciences Centre; Sunnybrook Research Institute; University of Toronto, Toronto, Canada
12. Baycrest Health Sciences, Rotman Research Institute, Toronto, Canada
13. Tanz Centre for Research in Neurodegenerative Diseases, University of Toronto, Toronto, Canada
14. Krembil Research Institute, University Health Network, Toronto, Canada
15. Department of Neurology, Erasmus Medical Centre, Rotterdam, Netherlands
16. Department of Neuroscience, Psychology, Drug Research and Child Health, University of Florence, Florence, Italy
17. Neuroradiology Unit, Department of Experimental and Clinical Biomedical Sciences, University of Florence, Florence, Italy
18. SOD Neurologia, Dipartimento di Neuroscienze degli Organi di Senso, AOU Careggi, Florence, Italy
19. Nuffield Department of Clinical Neurosciences, University of Oxford, Oxford, UK
20. Department of Neurobiology, Care Sciences and Society; Division of Clinical Geriatrics / Neurogeriatrics, Karolinska Institutet, Stockholm/Solna, Sweden
21. Theme Inflammation and Aging, Karolinska University Hospital, Sweden
22. Clinique Interdisciplinaire de Mémoire, CHU de Québec; CRCHU de Québec, Université Laval, Québec, Canada
23. Alzheimer's and Other Cognitive Disorders Unit, Hospital Clínic de Barcelona; FRCB-IDIBAPS; University of Barcelona; CIBERNED, Barcelona, Spain
24. Faculty of Medicine; Laboratory of Language Research, Centro de Estudos Egas Moniz, University of Lisbon, Lisbon, Portugal
25. Biogipuzkoa Health Research Institute, Neurosciences Area, Group of Neurodegenerative Diseases, San Sebastian, Spain
26. Cognitive Disorders Unit, Department of Neurology, Donostia University Hospital, San Sebastian, Spain
27. Division of Translational Genomics of Neurodegenerative Diseases, University of Tübingen, Germany
28. Department of Diagnostic and Interventional Neuroradiology, University of Tübingen, Germany
29. KU Leuven; University Hospitals Leuven; Department of Human Genetics; VIB Center for Molecular Neurology, Belgium
30. Douglas Research Centre, Department of Psychiatry, McGill University, Montreal, Canada
31. Montreal Neurological Institute, Department of Neurology & Neurosurgery, McGill University, Montreal, Canada
32. Reference Center for ALS, Département de Neurologie, AP-HP Hôpital Pitié-Salpêtrière, Paris, France
33. Paris Brain Institute (ICM), Sorbonne Université, Inserm U1127, CNRS UMR 7225, Paris, France
34. Neurologische Klinik, Ludwig-Maximilians-Universität München, Munich, Germany
35. Department of Neurology, University of Ulm, Ulm, Germany
36. Department of Neurosciences, Biomedical Research Institute BIOMED, Hasselt University, Belgium
37. VIB Center for Molecular Neurology; University of Antwerp; University Hospital of Antwerp (UZA), Belgium
38. Univ. Lille; Inserm; CHU Lille; Lille Neuroscience & Cognition; LabEx DISTALZ, Lille, France
39. University of Eastern Finland; Kuopio University Hospital, Kuopio, Finland
40. Kuopio University Hospital, Kuopio, Finland
41. University of Coimbra (multiple departments including Neurochemistry Laboratory; ICNAS; Neurology Department; Centre of Neurosciences and Cell Biology), Coimbra, Portugal
42. University of Leipzig; Max Planck Institute for Human Cognitive and Brain Sciences, Leipzig, Germany
43. University of Oulu; Oulu University Hospital; Research Unit of Clinical Medicine / Neurology, Finland
44. Department of Neurology, Charité Universitätsmedizin Berlin, Berlin, Germany
45. Center for Neurodegenerative Diseases and Aging Brain, University of Bari; Pia Fondazione Panico, Italy
46. Department of Neurology, Hospital de la Santa Creu i Sant Pau; FRCB-IDIBAPS; University of Barcelona; CIBERNED, Barcelona, Spain

### ALLFTD Investigators

Muhammad Abdullah[1]; Liana Apostolova[2]; Brian Appleby[3]; Sami Barmada[4]; Ece Bayram[5]; Bradley Boeve[6]; Hugo Botha[6]; Adam L. Boxer[7]; Andrea Bozoki[8,9,10]; Danielle Brushaber[11]; David Clark[2]; Ciaran M. Considine[12]; R. Ryan Darby[12]; Gregory S. Day[13]; Bradford Dickerson[3]; Dennis Dickson[13]; Kimiko Domoto-Reilly[14]; Emily Dwosh[15]; Kelley Faber[16]; Anne Fagan[17]; Julie A. Fields[18]; Jamie C. Fong[19]; Tatiana Foroud[20]; Leah K. Forsberg[6]; Douglas R. Galasko[21]; Ralitza Gavrilova[6,22]; Tania Gendron[13]; Daniel Geschwind[23]; Nupur Ghoshal[24]; Jill Goldman[25]; Jonathan Graff-Radford[6]; Neill Graff-Radford[13]; Ian M. Grant[26]; Murray Grossman[27]; Chadwick M. Hales[28]; Matthew Hall[7]; Hilary W. Heuer[7]; Lawrence S. Honig[29,30,31]; Ging-Yuek (Robin) Hsiung[15]; Eric Huang[7,32]; Edward D. Huey[29,30,31]; David Irwin[27]; Noah Johnson[11]; David T. Jones[6]; Kejal Kantarci[6]; David Knopman[6]; Tyler Kolander[6]; John Kornak[33]; Walter Kremers[11]; Justin Kwan[34]; Argentina Lario Lago[7]; Maria Lapid[18]; Shannon B. Lavigne[28]; Suzee Lee[7]; Gabriel C. Léger[21]; Irene Litvan[21]; Peter Ljubenkov[7]; Diane Lucente[35]; Ian R. Mackenzie[36]; Joseph C. Masdeu[37]; Lauren Massimo[27]; Scott McGinnis[35]; Corey T. McMillan[27]; Mario F. Mendez[38]; Carly Mester[11]; Toji Miyagawa[28]; Joie Molden[39]; Chiadi Onyike[40]; Alexander Pantelyat[41]; Belen Pascual[37]; Henry Paulson[4]; Leonard Petrucelli[13]; Peter Pressman[39]; Rosa Rademakers[42,43,44]; Vijay Ramanan[6]; Eliana Marisa Ramos[38]; Katherine P. Rankin[7]; Meghana Rao[6]; Katya Rascovsky[27]; Kristoffer W. Rhoads[14]; Aaron Ritter[45]; Erik D. Roberson[46]; Emily Rogalski[26]; Julio C. Rojas[7]; Howard J. Rosen[7]; Rodolfo Savica[6]; William Seeley[7]; Allison Snyder[34]; Adam M. Staffaroni[7]; Anna Campbell Sullivan[1]; Jeremy Syrjanen[11]; M. Carmela Tartaglia[47]; Jack Taylor[7]; Philip W. Tipton[13]; Marijne Vandebergh[42,43,44]; Sandra Weintraub[26]; Dylan Wint[45]; Bonnie Wong[35]; Jennifer Yokoyama[7]

1. Glenn Biggs Institute for Alzheimer's and Neurodegenerative Diseases, UT Health San Antonio
2. Department of Neurology, Indiana University, Indianapolis, IN, USA
3. Department of Neurology, Case Western Reserve University, Cleveland, OH, USA
4. Department of Neurology, University of Michigan, Ann Arbor, MI, USA
5. Parkinson and Other Movement Disorders Center, Department of Neurosciences, University of California San Diego, La Jolla, CA, USA
6. Department of Neurology, Mayo Clinic, Rochester, MN, USA
7. Department of Neurology, Memory and Aging Center, Weill Institute for Neurosciences, University of California, San Francisco, CA, USA
8. Department of Radiology, Michigan State University, East Lansing, MI, USA
9. Department of Neurology, Michigan State University, East Lansing, MI, USA
10. Department of Neurology, University of North Carolina, Chapel Hill, NC, USA
11. Department of Quantitative Health Sciences, Mayo Clinic, Rochester, MN, USA
12. Department of Neurology, Vanderbilt University, Nashville, TN, USA
13. Department of Neuroscience, Mayo Clinic, Jacksonville, FL, USA
14. Department of Neurology, University of Washington, Seattle, WA, USA
15. Division of Neurology, University of British Columbia, Vancouver, BC, Canada
16. Indiana University School of Medicine, National Centralized Repository for Alzheimer's, Indianapolis, IN, USA
17. Department of Neurology, Washington University School of Medicine, St. Louis, MO, USA
18. Department of Psychiatry and Psychology, Mayo Clinic, Rochester, MN, USA
19. UCSF Health Center for Clinical Genetics and Genomics, University of California, San Francisco, CA, USA
20. Department of Medical and Molecular Genetics, Indiana University School of Medicine, Indianapolis, IN, USA
21. Department of Neurosciences, University of California San Diego, La Jolla, CA, USA
22. Department of Clinical Genomics, Mayo Clinic, Rochester, MN, USA

23. Institute for Precision Health; Departments of Neurology, Psychiatry and Human Genetics, UCLA, Los Angeles, CA, USA
24. Departments of Neurology and Psychiatry, Washington University School of Medicine, St. Louis, MO, USA
25. Taub Institute for Research on Alzheimer's Disease and the Aging Brain, Department of Neurology, Columbia University Irving Medical Center, New York, NY, USA
26. Department of Psychiatry and Behavioral Sciences, Northwestern Feinberg School of Medicine, Chicago, IL, USA
27. Department of Neurology and Penn Frontotemporal Degeneration Center, Perelman School of Medicine, University of Pennsylvania, Philadelphia, PA, USA
28. Department of Neurology, Emory University School of Medicine, Atlanta, GA, USA
29. Taub Institute for Research on Alzheimer's Disease and the Aging Brain, Columbia University, New York, NY, USA
30. Department of Neurology, Columbia University, New York, NY, USA
31. Department of Psychiatry, Columbia University, New York, NY, USA
32. Department of Pathology, University of California San Francisco, San Francisco, CA, USA
33. Department of Epidemiology and Biostatistics, University of California, San Francisco, CA, USA
34. National Institute of Neurological Disorders and Stroke, USA
35. Department of Neurology, Massachusetts General Hospital and Harvard Medical School, Boston, MA, USA
36. Department of Pathology, University of British Columbia, Vancouver, BC, Canada
37. Department of Neurology, Houston Methodist, Houston, TX, USA
38. Department of Neurology, University of California, Los Angeles, CA, USA
39. Department of Neurology, University of Colorado, Aurora, CO, USA
40. Department of Psychiatry and Behavioral Sciences, Johns Hopkins University, Baltimore, MD, USA
41. Department of Neurology, Johns Hopkins University School of Medicine, Baltimore, MD, USA
42. VIB Center for Molecular Neurology, University of Antwerp, Antwerp, Belgium
43. Department of Biomedical Sciences, University of Antwerp, Antwerp, Belgium
44. Department of Neuroscience, Mayo Clinic, Jacksonville, FL, USA
45. Cleveland Clinic Lou Ruvo Center for Brain Health, Las Vegas, NV, USA
46. Department of Neurology, University of Alabama at Birmingham, Birmingham, AL, USA
47. Tanz Centre for Research in Neurodegenerative Diseases, Division of Neurology, University of Toronto, Toronto, ON, Canada

### Participant Demographics

#### eTable 1a: Demographic characteristics of GENFI participants

| Characteristic | All | | | *C9orf72* | | | *GRN* | | | *MAPT* | | |
|---|---|---|---|---|---|---|---|---|---|---|---|---|
| | Total | Carriers | Non-carriers | Carriers | Non-carriers | Between group comparison | Carriers | Non-carriers | Between group comparison | Carriers | Non-carriers | Between group comparison |
| Sample size (*n*) | 154 | 85 | 69 | 41 | 29 | NA | 25 | 21 | NA | 19 | 19 | NA |
| Number of visits (mean ± SD) | 1.8 ± 1.2 | 1.7 ± 1.0 | 1.9 ± 1.4 | 1.5 ± 0.9 | 1.6 ± 0.9 | $t=-0.41$, $p=0.68$, $d=-0.10$ | 1.8 ± 1.1 | 2.4 ± 2.2 | $t=-1.18$, $p=0.25$, $d=-0.37$ | 1.9 ± 1.0 | 1.9 ± 1.0 | $t=0$, $p=1$, $d=0$ |
| Time between visits, years (mean ± SD) | 1.55 ± 0.75 | 1.47 ± 0.65 | 1.61 ± 0.82 | 1.50 ± 0.56 | 2.21 ± 1.25 | $t=0.41$, $p=0.68$, $d=-0.10$ | 1.34 ± 0.47 | 1.53 ± 0.49 | $t=-1.33$, $p=0.19$, $d=-0.40$ | 1.56 ± 0.89 | 1.18 ± 0.26 | |
| Age at visit, years (mean ± SD) | 26.1 ± 2.9 | 26.1 ± 3.0 | 26.0 ± 2.8 | 26.7 ± 2.9 | 26.5 ± 2.5 | $t=0.35$, $p=0.72$, $d=0.07$ | 25.8 ± 2.8 | 26.1 ± 2.7 | $t=-0.53$, $p=0.60$, $d=-0.11$ | 25.6 ± 3.2 | 25.5 ± 3.0 | $t=0.15$, $p=0.88$, $d=0.04$ |
| Sex (*n*) female / male | <100 / <100 | <50 / >30 | >30 / >30 | NR | NR | $x^2=0$, $p=1$, $V=0$ | NR | NR | $x^2=0.26$, $p=0.61$, $V=0.05$ | NR | NR | $x^2=0$, $p=1$, $V=0$ |
| Education, years (mean ± SD) | 14.7 ± 2.3 | 15.0 ± 2.1 | 14.3 ± 2.4 | 15.0 ± 2.4 | 14.5 ± 2.5 | $t=1.07$, $p=0.29$, $d=0.21$ | 14.6 ± 1.8 | 14.6 ± 2.7 | $t=-0.43$, $p=-0.67$, $d=-0.09$ | 15.2 ± 1.9 | 13.7 ± 1.9 | $t=3.40$, $p=0.001$**, $d=0.79$ |
| Handedness (*n*)<br>Right<br>Left<br>Ambidextrous | <br>50-100<br><50<br><10 | <br>50-100<br>10-30<br><10 | <br>50-100<br><10<br><10 | <br>30-50<br><10<br><10 | <br><30<br><10<br><10 | $x^2=1.34$, $p=0.25$, $V=0.11$ | <br><30<br><10<br><10 | <br><30<br><10<br><10 | $x^2=1.49$, $p=0.22$, $V=0.13$ | <br><30<br><10<br><10 | <br><30<br><10<br><10 | $x^2=3.36$, $p=0.07$, $V=0.21$ |
| Sites (*n*) | 25 | 23 | 22 | 16 | 18 | NA | 13 | 8 | NA | 10 | 7 | NA |

#### eTable 1b: Demographic characteristics of ALLFTD participants

| Characteristic | All | | | *C9orf72* | | | *GRN* | | | *MAPT* | | |
|---|---|---|---|---|---|---|---|---|---|---|---|---|
| | Total | Carriers | Non-carriers | Carriers | Non-carriers | Between group comparison | Carriers | Non-carriers | Between group comparison | Carriers | Non-carriers | Between group comparison |
| Sample size (*n*) | >100 | >50 | >50 | 31 | 19 | NA | <20 | <10 | NA | 21 | 20 | NA |
| Number of visits (mean ± SD) | 2.7 ± 2.0 | 2.9 ± 2.2 | 2.3 ± 1.7 | 2.4 ± 2.1 | 1.7 ± 0.9 | $t=1.60$, $p=0.12$, $d=0.39$ | 2.3 ± 1.8 | 2.2 ± 0.8 | $t=0.15$, $p=0.89$, $d=0.06$ | 2.9 ± 2.2 | 2.9 ± 2.2 | $t=1.33$, $p=0.19$, $d=0.42$ |
| Time between visits, years (mean ± SD) | 1.25 ± 0.50 | 1.28 ± 0.56 | 1.18 ± 0.35 | 1.17 ± 0.29 | 1.24 ± 0.31 | $t=-0.79$, $p=0.43$, $d=-0.24$ | 1.24 ± 0.49 | 1.22 ± 0.28 | $t=0.10$, $p=0.92$, $d=0.05$ | 1.37 ± 0.70 | 1.15 ± 0.38 | $t=1.26$, $p=0.22$, $d=0.39$ |
| Age at visit, years (mean ± SD) | 25.8 ± 3.5 | 25.7 ± 3.4 | 25.9 ± 3.5 | 26.1 ± 3.3 | 27.8 ± 2.3 | $t=-2.66$, $p=0.009$**, $d=-0.55$ | NR | NR | $t=-0.18$, $p=0.86$, $d=-0.07$ | 24.8 ± 3.6 | 24.6 ± 3.7 | $t=0.40$, $p=0.69$, $d=0.07$ |
| Sex (*n*) female / male | >50 / <50 | >30 / >20 | >20 / <20 | NR | NR | $x^2=5.28e-31$, $p=1$, $V=6.99e-17$ | NR | NR | $x^2=0.05$, $p=0.82$, $V=0.04$ | NR | NR | $x^2=0.04$, $p=0.85$, $V=0.02$ |
| Education, years (mean ± SD) | 15.3 ± 2.3 | 15.3 ± 2.2 | 15.3 ± 2.4 | 15.6 ± 2.1 | 15.6 ± 2.6 | $t=-0.68$, $p=0.50$, $d=-0.14$ | NR | NR | $t=1.49$, $p=0.15$, $d=0.54$ | 14.3 ± 2.1 | 14.9 ± 2.5 | $t=-1.17$, $p=0.24$, $d=-0.20$ |
| Handedness (*n*)<br>Right<br>Left<br>Ambidextrous | <br><100<br>10-50<br><10 | <br>50-100<br><10<br><10 | <br>30-50<br><10<br><10 | <br>10-30<br><10<br><10 | <br>10-30<br><10<br><10 | $x^2=12.03$, $p=0.002$**, $V=0.33$ | <br><10<br><10<br><10 | <br><10<br><10<br><10 | $x^2=4.24$, $p=0.04$, $V=0.35$ | <br>10-30<br><10<br><10 | <br>10-30<br><10<br><10 | $x^2=0.17$, $p=0.68$, $V=0.04$ |
| Sites (*n*) | 13 | 12 | 9 | 11 | 8 | NA | 6 | 3 | NA | 6 | 7 | NA |

*p<0.05, **p<0.01 between carriers and non-carriers. Some participant counts and all handedness counts are reported in ranges to prevent unblinding participants who may be left-handed or ambidextrous. Abbreviations: SD, standard deviation; NA, not applicable; NR, not reported due to risk of unblinding.

### Neuroimaging

#### Summary of regions of interest (ROIs) assessed

Preprocessing was conducted in FreeSurfer v7.4.1, and included N3 bias-field correction and intensity normalization, skull stripping using watershed segmentation, affine Talairach registration, white matter and pial surface reconstruction with spherical surface registration. Cortical parcellation was conducted using the Desikan-Killiany atlas, and subcortical volumetric segmentation using FreeSurfer's probabilistic atlas, Aseg.

- **Common volumetric ROIs across *C9orf72*, *GRN*, and *MAPT*:** total brain, medial orbitofrontal cortex (mOFC), frontal pole
- **Specific ROIs for *C9orf72*:** thalamus, insula, rostral anterior cingulate (rACC)
- **Specific ROIs for *GRN*:** putamen, insula, rostral anterior cingulate (rACC)
- **Specific ROIs for *MAPT*:** anterior medial temporal lobe (hippocampus, amygdala, entorhinal cortex, parahippocampus, temporal pole)

#### Post-hoc statistics for *C9orf72* thalamus and rACC

**Thalamus:**

Post-hoc models, where the left and right thalamus were fitted separately, revealed consistent patterns, with slightly greater effect sizes in the right than the left. Compared to non-carriers, young adult *C9orf72* carriers had smaller left thalamic volumes ($b$=-463.3mm$^3$, SE=171.7, $p$=0.012, $f^2$=0.26), males had larger left thalami than females ($b$=825.659mm$^3$, SE=174.0, $p$=0.0001, $f^2$=0.77), and no group-by-age interaction was observed. In the right thalamus, young adult *C9orf72* carriers had smaller volumes compared to non-carriers ($b$=-527.8mm$^3$, SE=154.4, $p$=0.0021, $f^2$=0.35), and males had larger volumes than females ($b$=727.5 mm$^3$, SE=154.5, $p$=0.0001, $f^2$=0.58). The main effect of age ($b$=-56.0 mm$^3$, SE=30.2, $p$=0.068, $f^2$=0.08) and group-by-age interaction ($b$=-68.6mm$^3$, SE=36.2, $p$=0.0620, $f^2$=0.05) on right thalamic volumes trended towards significance, where non-carriers showed age-related decline ($b$=-56.0mm$^3$/year, 95% CI [-116, 4.31]) and carriers had relatively stable trajectories ($b$=12.6mm$^3$/year, 95% CI [-24, 49.22]).

**Rostral Anterior Cingulate Cortex (rACC):**

Post-hoc models, where the left and right rACC were fitted separately, demonstrated a significant group-by-age interaction in the left rACC ($b$=52.0mm$^3$, SE=18.8, $p$=0.0072, $f^2$=0.11), with a similar pattern of non-carriers showing age-related decline ($b$=-58.3mm$^3$/year, 95% CI [-89.5, -27.2]) compared to stable trajectories in carriers ($b$=-6.3mm$^3$/year, 95% CI [-25.8, 13.2]). No significant group-by-age interaction or main effects were observed in the right rACC (p>0.05).

#### eTable 2. Linear mixed-effects model main outcomes for *GRN* regions of interest.

| ***GRN* model** | **Group main effect** | **Age main effect** | **Group x Age interaction** |
|---|---|---|---|
| Total brain | $b$=19067.1, SE=25503.3, $p$=0.47, $f^2$=0.03 | $b$=-1103.9, SE=1479.8, $p$=0.46, $f^2$=0.03 | $b$=-215.7, SE=2068.9, $p$=0.92, $f^2$=2.2e-4 |
| Frontal pole | $b$=31.3, SE=58.7, $p$=0.60, $f^2$=0.05 | $b$=-17.9, SE=11.4, $p$=0.12, $f^2$=0.31 | $b$=11.06, SE=15.2, $p$=0.47, $f^2$=0.01 |
| mOFC | $b$=396.8, SE=322.5, $p$=0.24, $f^2$=0.10 | $b$=-54.4, SE=36.8, $p$=0.15, $f^2$=0.09 | $b$=16.5, SE=50.7, $p$=0.75, $f^2$=2.2e-3 |
| rACC | $b$=248.1, SE=217.8, $p$=0.27, $f^2$=0.05 | $b$=248.1, SE=217.8, $p$=0.13, $f^2$=3.5e-4 | $b$=26.2, SE=25.4, $p$=0.31, $f^2$=0.02 |
| Insula | $b$=159.3, SE=316.5, $p$=0.62, $f^2$=0.03 | $b$=82.3, SE=37.5, $p$=0.03, $f^2$=0.03 | $b$=1.1, SE=52.1, p=0.98, $f^2$=9.3e-6 |
| Putamen | $b$=12.9, SE=236.9, $p$=0.96, $f^2$=0.02 | $b$=22.3, SE=30.3, $p$=0.47, $f^2$=0.14 | $b$=-23.4, SE=41.6, $p$=0.58, $f^2$=6.4e-3 |

All volumes are measured in $mm^3$. Abbreviations: mOFC = medial orbitofrontal cortex; rACC = rostral anterior cingulate cortex.

#### eTable 3. Linear mixed-effects model main outcomes for *MAPT* regions of interest.

| ***MAPT* model** | **Group main effect** | **Age main effect** | **Group x Age interaction** |
|---|---|---|---|
| Total brain | $b$=-6509.8, SE=24331.7, $p$=0.79, $f^2$=0.02 | $b$=-281.8, SE=1171.9, $p$=0.81, $f^2$=0.07 | $b$=-3049.8, SE=1267.7, *$p$=0.02, $f^2$=0.06 |
| mOFC | $b$=7.8, SE=268.1, $p$=0.98, $f^2$=1.4e-3 | $b$=-16.2, SE=28.0, $p$=0.57, $f^2$=0.10 | $b$=-57.0, SE=31.6, $p$=0.07, $f^2$=0.03 |
| rACC | $b$=-75.9, SE=232.6, $p$=0.75, $f^2$=5.4e-3 | $b$=-40.5, SE=17.6, *$p$=0.02, $f^2$=0.01 | $b$=15.6, SE=19.3, $p$=0.42, $f^2$=6.7e-3 |
| Anterior medial temporal pole | $b$=-125.7, SE=512.7, $p$=0.81, $f^2$=8.5e-3 | $b$=-5.1, SE=53.8, $p$=0.92, $f^2$=4.0e-3 | $b$=-58.3, SE=60.6, $p$=0.34, $f^2$=9.5e-3 |

All volumes are measured in $mm^3$. *$p$<0.05. Abbreviations: mOFC = medial orbitofrontal cortex; rACC = rostral anterior cingulate cortex.

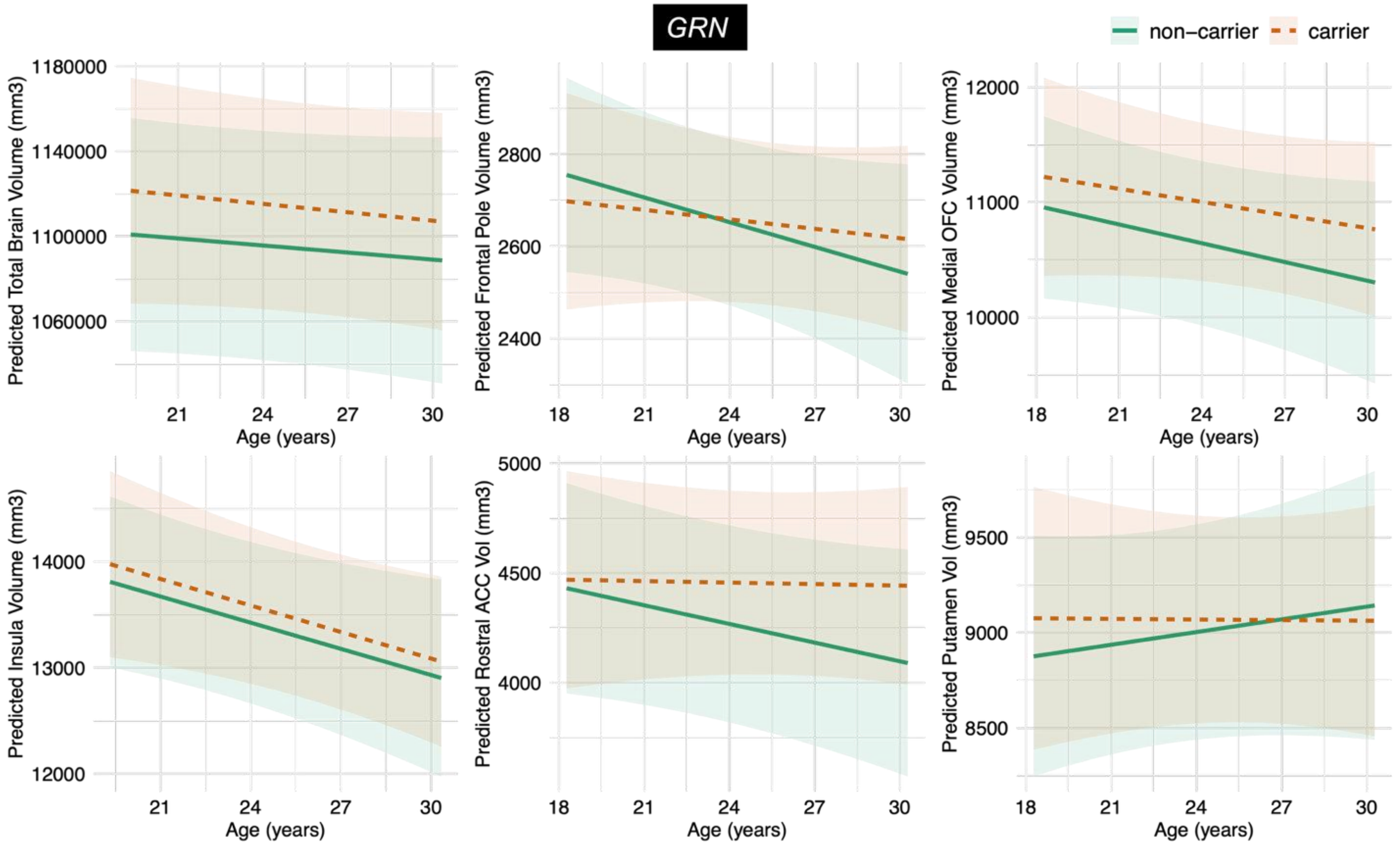

**eFigure 1. Predicted trajectories of young adult *GRN* carriers compared to non-carriers.** No group main effects or longitudinal trajectory differences were observed for all regions of interest. The plotted confidence intervals reflect pointwise uncertainty in the predicted marginal means for each group and do not represent uncertainty in the group-by-age interaction term, which tests differences in age-related slopes.

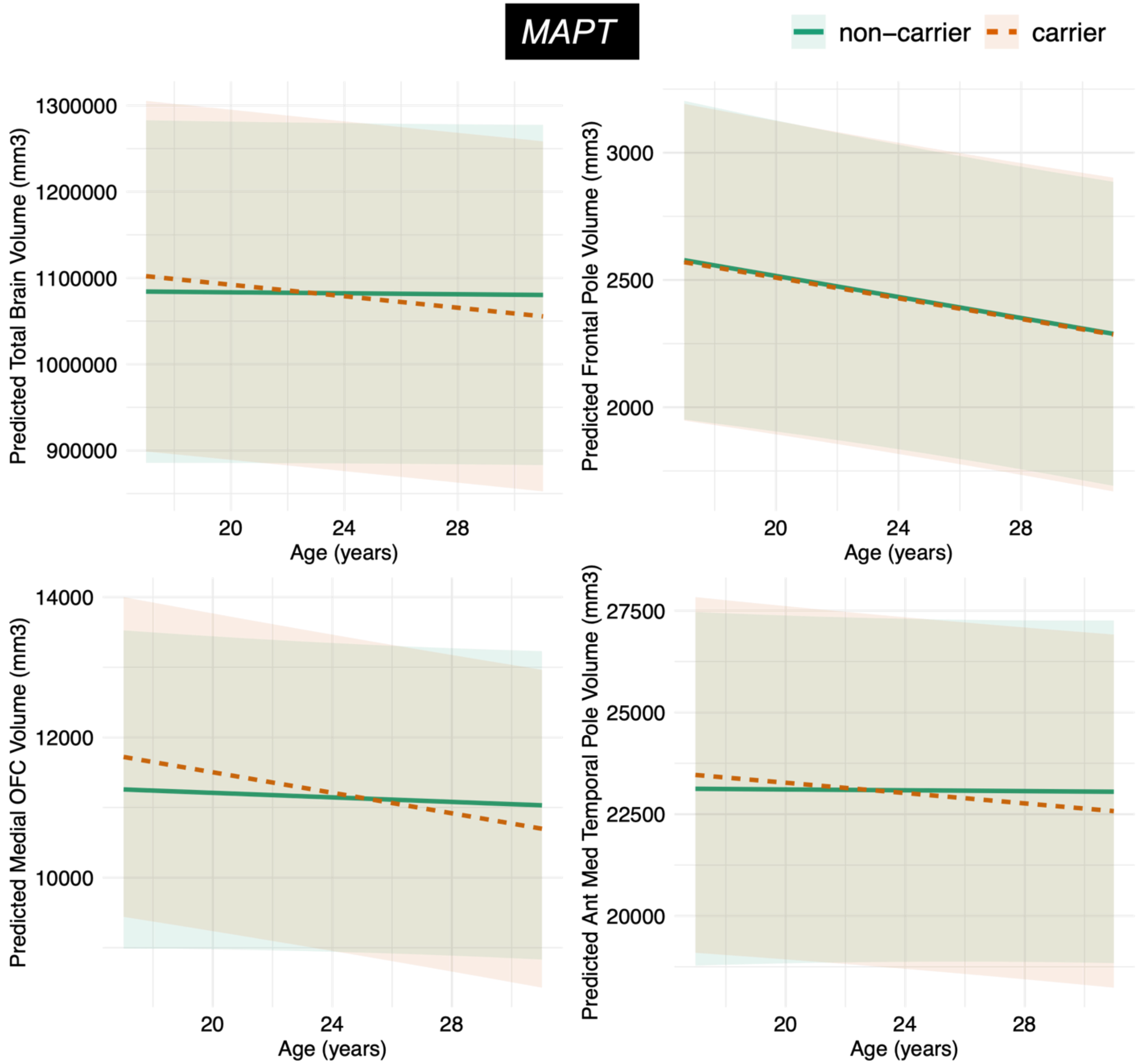

**eFigure 2. Predicted trajectories of young adult *MAPT* carriers compared to non-carriers.** A statistically significant interaction was observed for the total brain, and trended towards significance in the mOFC, where carriers demonstrated faster rates of age-related declines across age 18 to 30 compared to relatively stable trajectories in non-carriers. No group main effects or interactions were observed for the frontal pole and anterior medial temporal pole. The plotted confidence intervals reflect pointwise uncertainty in the predicted marginal means for each group and do not represent uncertainty in the group-by-age interaction term, which tests differences in age-related slopes.

#### Linear-mixed effects model outcomes from normative modelling-derived deviation z-scores

##### eTable 4a: Normative modelling analysis outcomes for *C9orf72.*

| Measure | Sex | ROI | Model Outcomes |
|---|---|---|---|
| Volume | M | Left thalamus | Group: *b*=0.98, SE=3.50, *p*=0.78, $f^2$=0.02<br>Group-by-age: *b*=-0.02, SE=0.13, *p*=0.87, $f^2$=1.28e-3 |
| | | Right thalamus | Group: *b*=-7.53, SE=2.91, **p*=0.013, $f^2$=0.07<br>Group-by-age: *b*=0.25, SE=0.11, **p*=0.026, $f^2$=0.25<br>Carrier slope: *b*=0.15, 95% CI [0.035, 0.2675]<br>Non-carrier slope: *b*=-0.10, 95% CI [-0.29, 0.09] |
| | F | Left thalamus | Group: *b*=-0.46, SE=2.53, *p*=0.86, $f^2$=0.02<br>Group-by-age: *b*=0.03, SE=0.09, *p*=0.75, $f^2$=2.11e-3 |
| | | Right thalamus | Group: *b*=4.52, SE=2.53, *p*=0.079, $f^2$=0.05<br>Group-by-age: *b*=-0.15, SE=0.09, *p*=0.116, $f^2$=0.05 |
| Cortical thickness | M | Left insula | Group: *b*=-2.59, SE=2.79, *p*=0.36, $f^2$=6.84e-8<br>Group-by-age: *b*=0.09, SE=0.10, *p*=0.36, $f^2$=0.04 |
| | | Right insula | Group: *b*=0.54, SE=2.89, *p*=0.85, $f^2$=8.09e-4<br>Group-by-age: *b*=-0.02, SE=0.10, *p*=0.83, $f^2$=2.09e-3 |
| | F | Left insula | Group: *b*=-0.76, SE=1.83, *p*=0.68, $f^2$=6.73e-4<br>Group-by-age: *b*=0.03, SE=0.07, *p*=0.66, $f^2$=3.69e-3 |
| | | Right insula | Group: *b*=-1.79, SE=1.71, *p*=0.30, $f^2$=7.31e-3<br>Group-by-age: *b*=0.07, SE=0.06, *p*=0.27, $f^2$=0.03 |
| Cortical thickness | M | Left frontal pole | Group: *b*=-2.63, SE=1.92, *p*=0.18, $f^2$=2.52e-3<br>Group-by-age: *b*=0.09, SE=0.07, *p*=0.19, $f^2$=0.08 |
| | | Right frontal pole | Group: *b*=-2.52, SE=1.85, *p*=0.18, $f^2$=0.02<br>Group-by-age: *b*=0.10, SE=0.07, *p*=0.15, $f^2$=0.10 |
| | F | Left frontal pole | Group: *b*=-1.00, SE=1.68, *p*=0.56, $f^2$=1.32e-3<br>Group-by-age: *b*=0.04, SE=0.06, *p*=0.57, $f^2$=7.15e-3 |
| | | Right frontal pole | Group: *b*=-0.22, SE=1.62, *p*=0.89, $f^2$=0.01<br>Group-by-age: *b*=0.02, SE=0.06, *p*=0.78, $f^2$=1.77e-3 |
| Cortical thickness | M | Left mOFC | Group: *b*=-5.08, SE=2.43, **p*=0.043, $f^2$=0.008<br>Group-by-age: *b*=0.18, SE=0.09, *p*=0.055, $f^2$=0.18 |
| | | Right mOFC | Group: *b*=4.88, SE=2.60, *p*=0.068, $f^2$=0.009<br>Group-by-age: *b*=-0.19, SE=0.09, p=0.060, $f^2$=0.17 |
| | F | Left mOFC | Group: *b*=-0.58, SE=1.50, *p*=0.70, $f^2$=8.68e-3<br>Group-by-age: b=0.01, SE=0.06, *p*=0.79, $f^2$=1.59e-3 |
| | | Right mOFC | Group: *b*=-0.60, SE=2.05, *p*=0.77, $f^2$=5.99e-4<br>Group-by-age: *b*=0.02, SE=0.08, *p*=0.79, $f^2$=1.62e-3 |
| Cortical thickness | M | Left rACC | Group: *b*=-3.27, SE=2.21, *p*=0.15, $f^2$=0.01<br>Group-by-age: *b*=0.11, SE=0.08, *p*=0.17, $f^2$=0.09 |
| | | Right rACC | Group: *b*=-4.47, SE=2.61, *p*=0.09, $f^2$=5.26e-3<br>Group-by-age: *b*=0.16, SE=0.09, *p*=0.11, $f^2$=0.12 |
| | F | Left rACC | Group: *b*=0.95, SE=1.62, *p*=0.56, $f^2$=3.31e-3<br>Group-by-age: *b*=-0.03, SE=0.06, *p*=0.59, $f^2$=6.58e-3 |
| | | Right rACC | Group: *b*=3.41, SE=1.90, *p*=0.08, $f^2$=0.03<br>Group-by-age: *b*=-0.12, SE=0.07, *p*=0.11, $f^2$=0.06 |

Abbreviations: mOFC=medial orbitofrontal cortex; rACC=rostral anterior cingulate cortex.

#### eTable 4b: Normative modelling analysis outcomes for *GRN.*

| Measure | Sex | ROI | Model Outcomes |
|---|---|---|---|
| Volume | M | Left putamen | Group: *b*=-2.99, SE=2.76, *p*=0.29, $f^2$=3.39e-3<br>Group-by-age: *b*=0.02, SE=0.08, *p*=0.29, $f^2$=0.05 |
| | | Right putamen | Group: *b*=-0.83, SE=0.93, *p*=0.38, $f^2$=4.70e-3<br>Group-by-age: *b*=0.04, SE=0.03, *p*=0.26, $f^2$=0.05 |
| | F | Left putamen | Group: *b*=-3.14, SE=2.19, *p*=0.16, $f^2$=0.07<br>Group-by-age: *b*=0.13, SE=0.08, *p*=0.13, $f^2$=0.10 |
| | | Right putamen | Group: *b*=-1.16, SE=1.84, *p*=0.53, $f^2$=0.03<br>Group-by-age: *b*=0.05, SE=0.07, *p*=0.49, $f^2$=0.02 |
| Cortical thickness | M | Left insula | Group: *b*=-3.21, SE=2.25, *p*=0.17, $f^2$=0.01<br>Group-by-age: *b*=0.13, SE=0.08, *p*=0.14, $f^2$=0.09 |
| | | Right insula | Group: *b*=5.24, SE=2.09, **p*=0.020, $f^2$=0.05<br>Group-by-age: *b*=-0.18, SE=0.08, **p*=0.026, $f^2$=0.23<br>Carrier slope: *b*=-0.06, 95% CI [-0.19, 0.06]<br>Non-carrier slope: *b*=0.12, 95% CI [0.019, 0.225] |
| | F | Left insula | Group: *b*=-7.32, SE=2.74, **p*=0.012, $f^2$=0.04<br>Group-by-age: *b*=0.26, SE=0.10, **p*=0.018, $f^2$=0.26<br>Carrier slope: *b*=0.09, 95% CI [-0.07, 0.24]<br>Non-carrier slope: *b*=-0.17, 95% CI [-0.32, -0.027] |
| | | Right insula | Group: *b*=0.58, SE=1.93, *p*=0.77, $f^2$=2.37e-3<br>Group-by-age: *b*=-0.03, SE=0.07, *p*=0.73, $f^2$=4.94e-3 |
| Cortical thickness | M | Left frontal pole | Group: *b*=-5.07, SE=1.63, ***p*=0.005, $f^2$=0.01<br>Group-by-age: *b*=0.19, SE=0.06, ***p*=0.005, $f^2$=0.38<br>Carrier slope: b=0.08, 95% CI [-0.02, 0.17]<br>Non-carrier slope: *b*=-0.11, 95% CI [-0.19, -0.03] |
| | | Right frontal pole | Group: *b*=1.73, SE=1.55, *p*=0.28, $f^2$=0.10<br>Group-by-age: *b*=-0.05, SE=0.06, *p*=0.39, $f^2$=0.03 |
| | F | Left frontal pole | Group: *b*=1.74, SE=2.14, *p*=0.42, $f^2$=0.15<br>Group-by-age: *b*=-0.09, SE=0.08, *p*=0.29, $f^2$=0.05 |
| | | Right frontal pole | Group: *b*=-2.70, SE=1.89, *p*=0.17, $f^2$=0.11<br>Group-by-age: *b*=0.07, SE=0.07, *p*=0.32, $f^2$=0.04 |
| Cortical thickness | M | Left mOFC | Group: *b*=-4.59, SE=2.00, **p*=0.031, $f^2$=0.03<br>Group-by-age: *b*=-0.17, SE=0.075, **p*=0.036, $f^2$=0.20<br>Carrier slope: *b*=-0.09, 95% CI [-0.21, 0.018]<br>Non-carrier slope: *b*=0.07, 95% CI [-0.035, 0.18] |
| | | Right mOFC | Group: *b*=-2.98, SE=1.72, p=0.10, $f^2$=0.10<br>Group-by-age: *b*=0.09, SE=0.09, p=0.29, $f^2$=0.05 |
| | F | Left mOFC | Group: *b*=-3.05, SE=1.87, *p*=0.11, $f^2$=0.03<br>Group-by-age: *b*=-0.12, SE=0.07, *p*=0.088, $f^2$=0.20 |
| | | Right mOFC | Group: *b*=0.94, SE=2.30, *p*=0.69, $f^2$=0.17<br>Group-by-age: *b*=0.01, SE=0.09, *p*=0.91, $f^2$=5.65e-4 |
| Cortical thickness | M | Left rACC | Group: *b*=-3.00, SE=1.86, *p*=0.12, $f^2$=0.03<br>Group-by-age: *b*=0.10, SE=0.07, *p*=0.15, $f^2$=0.09 |
| | | Right rACC | Group: *b*=0.49, SE=2.33, *p*=0.84, $f^2$=0.03<br>Group-by-age: *b*=-0.03, SE=0.09, *p*=0.73, $f^2$=4.87e-3 |
| | F | Left rACC | Group: *b*=-2.75, SE=.90, *p*=0.16, $f^2$=0.02<br>Group-by-age: *b*=0. 111, SE=0.07, *p*=0.14, $f^2$=0.09 |
| | | Right rACC | Group: *b*=4.95, SE=2.09, **p*=0.03, $f^2$=0.01<br>Group-by-age: *b*=-0.18, SE=0.08, **p*=0.03, $f^2$=0.22 |

Abbreviations: mOFC=medial orbitofrontal cortex; rACC=rostral anterior cingulate cortex.

#### eTable 4c: Normative modelling analysis outcomes for *MAPT.*

| Measure | Sex | ROI | Model Outcomes |
|---|---|---|---|
| Volume | M | Left hippocampus | Group: *b*=-2.58, SE=1.09, **p*=0.024, $f^2$=0.01<br>Group-by-age: *b*=0.11, SE=0.04, **p*=0.012, $f^2$=0.20<br>Carrier slope: *b*=0.07, 95% CI [0.04, 0.11]<br>Non-carrier slope: *b*=-0.03, 95% CI [-0.11, 0.04] |
| | | Right hippocampus | Group: *b*=1.08, SE=1.57, *p*=0.50, $f^2$=0.01<br>Group-by-age: *b*=-0.05, SE=0.06, *p*=0.43, $f^2$=0.20 |
| | F | Left hippocampus | Group: *b*=1.75, SE=0.85, **p*=0.047, $f^2$=0.04<br>Group-by-age: *b*=-0.08, SE=0.03, **p*=0.017, $f^2$=0.09<br>Carrier slope: *b*=0.021, 95% CI [-0.02, 0.06]<br>Non-carrier slope: *b*=0.10, 95% CI [0.050, 0.153] |
| | | Right hippocampus | Group: *b*=-1.45, SE=0.89, *p*=0.11, $f^2$=0.06<br>Group-by-age: *b*=-0.07, SE=0.03, **p*=0.041, $f^2$=0.06<br>Carrier slope: *b*=0.02, 95% CI [-0.02, 0.06]<br>Non-carrier slope: *b*=0.09, 95% CI [0.04, 0.14] |
| Volume | M | Left amygdala | Group: *b*=3.27, SE=2.42, *p*=0.19, $f^2$=1.40e-4<br>Group-by-age: *b*=0.08, SE=0.05, *p*=0.08, $f^2$=0.05 |
| | | Right amygdala | Group: *b*=1.11, SE=1.94, *p*=0.57, $f^2$=1.45e-3<br>Group-by-age: *b*=-0.04, SE=0.07, *p*=0.55, $f^2$=0.01 |
| | F | Left amygdala | Group: *b*=2.81, SE=1.30, **p*=0.037, $f^2$=0.04<br>Group-by-age: *b*=-0.13, SE=0.05, **p*=0.016, $f^2$=0.09<br>Carrier slope: *b*=0.01, 95% CI [-0.052, 0.075]<br>Non-carrier slope: *b*=0.137, 95% CI [0.06, 0.22] |
| | | Right amygdala | Group: *b*=0.64, SE=1.17, *p*=0.59, $f^2$=0.04<br>Group-by-age: *b*=-0.01, SE=0.05, *p*=0.76, $f^2$=1.41e-3 |
| Cortical thickness | M | Left frontal pole | Group: *b*=-0.78, SE=1.24, *p*=0.53, $f^2$=1.66e-3<br>Group-by-age: *b*=0.03, SE=0.05, *p*=0.46, $f^2$=0.02 |
| | | Right frontal pole | Group: *b*=2.96, SE=1.32, **p*=0.03, $f^2$=0.36<br>Group-by-age: *b*=-0.08, SE=0.05, *p*=0.11, $f^2$=0.09 |
| | F | Left frontal pole | Group: *b*=-3.55, SE=1.38, **p*=0.014, $f^2$=2.90e-5<br>Group-by-age: *b*=0.14, SE=0.05, **p*=0.011, $f^2$=0.10<br>Carrier slope: *b*=-0.004, 95% CI [-0.07, 0.06]<br>Non-carrier slope: *b*=-0.147, 95% CI [-0.23, -0.06] |
| | | Right frontal pole | Group: *b*=0.36, SE=1.21, *p*=0.77, $f^2$=2.61e-4<br>Group-by-age: *b*=-0.01, SE=0.05, *p*=0.79, $f^2$=1.05e-3 |
| Cortical thickness | M | Left temporal pole | Group: *b*=-3.12, SE=2.06, *p*=0.14, $f^2$=0.04<br>Group-by-age: *b*=0.11, SE=0.08, *p*=0.17, $f^2$=0.06 |
| | | Right temporal pole | Group: *b*=3.82, SE=3.15, *p*=0.085, $f^2$=2.72e-3<br>Group-by-age: *b*=0.14, SE=0.08, *p*=0.095, $f^2$=0.09 |
| | F | Left temporal pole | Group: *b*=-0.60, SE=1.63, *p*=0.71, $f^2$=2.61e-4<br>Group-by-age: *b*=0.04, SE=0.07, *p*=0.56, $f^2$=1.05e-3 |
| | | Right temporal pole | Group: *b*=-0.16, SE=1.70, *p*=0.93, $f^2$=0.02<br>Group-by-age: *b*=-0.004, SE=0.07, *p*=0.95, $f^2$=4.96e-5 |
| Cortical thickness | M | Left mOFC | Group: *b*=0.22, SE=1.83, *p*=0.91, $f^2$=0.18<br>Group-by-age: *b*=0.02, SE=0.07, *p*=0.80, $f^2$=1.99e-3 |
| | | Right mOFC | Group: *b*=-1.52, SE=1.85, *p*=0.42, $f^2$=0.03<br>Group-by-age: *b*=0.07, SE=0.07, *p*=0.30, $f^2$=0.03 |
| | F | Left mOFC | Group: *b*=1.64, SE=1.18, *p*=0.17, $f^2$=1.48e-3<br>Group-by-age: *b*=-0.07, SE=0.05, *p*=0.15, $f^2$=0.03 |
| | | Right mOFC | Group: *b*=1.22, SE=1.17, *p*=0.31, $f^2$=7.38e-3<br>Group-by-age: *b*=-0.05, SE=0.05, *p*=0.25, $f^2$=0.02 |

Abbreviations: mOFC=medial orbitofrontal cortex.

### Null-hypothesis Testing for Executive Function and Plasma Biomarker Outcomes

#### eTable 5: Bayes Factors for the Group-By-Age Interaction in Executive Function and Plasma Biomarker Models

| Outcome | | $BF_{01}$ (evidence for the null) |
|---|---|---|
| *C9orf72* | Executive function composite | 2.87 |
| | Plasma NfL | 2.26 |
| | Plasma GFAP | 0.53 |
| GRN | Executive function composite | 2.90 |
| | Plasma NfL | 1.70 |
| | Plasma GFAP | 1.26 |
| *MAPT* | Executive function composite | 2.81 |
| | Plasma NfL | 1.66 |
| | Plasma GFAP | 2.08 |

### Behavioural Outcomes

#### eTable 6: Behavioural Outcome Statistics

| Model | | Chi-square statistics |
|---|---|---|
| *C9orf72* | Disinhibition | $\chi^2$=2.28e-3, *p*=1 |
| | Apathy | $\chi^2$=2.49e-31, p=1 |
| | Empathy | $\chi^2$=0, p=1 |
| | Obsessive-compulsive disorder | $\chi^2$=0, p=1 |
| | Appetite changes | $\chi^2$=0.54, p=0.46 |
| | Delusions | $\chi^2$=3.70e-30, p=1 |
| | Hallucinations | $\chi^2$=2.28e-30, p=1 |
| | Depression | $\chi^2$=0.69, p=0.41 |
| | Anxiety | $\chi^2$=2.64, p=0.10 |
| | Aggression | $\chi^2$=0.21, p=0.65 |
| GRN | Disinhibition | $\chi^2$=8.49e-32, *p*=1 |
| | Apathy | $\chi^2$=2.37e-30, p=1 |
| | Empathy | $\chi^2$=1.43e-30, p=1 |
| | Obsessive-compulsive disorder | $\chi^2$=0, p=1 |
| | Appetite changes | $\chi^2$=2.37e-30, p=1 |
| | Delusions | $\chi^2$=4.07e-33, p=1 |
| | Hallucinations | $\chi^2$=4.07-e33, p=1 |
| | Depression | $\chi^2$=3.67e-4, p=0.98 |
| | Anxiety | $\chi^2$=0.37, p=0.55 |
| | Aggression | $\chi^2$=1.11e-31, p=1 |
| *MAPT* | Disinhibition | $\chi^2$=1.15e-29, *p*=1 |
| | Apathy | $\chi^2$=4.52e-31, p=1 |
| | Empathy | $\chi^2$=1.23e-31, p=1 |
| | Obsessive-compulsive disorder | $\chi^2$=2.93e-32, p=1 |
| | Appetite changes | $\chi^2$=0.68, p=0.41 |
| | Delusions | $\chi^2$=1.20e-29, p=1 |
| | Hallucinations | $\chi^2$=1.53e-29, p=1 |
| | Depression | $\chi^2$=0.28, p=0.59 |
| | Anxiety | $\chi^2$=1.63, p=0.20 |
| | Aggression | $\chi^2$=0.01, p=92 |

### Sensitivity Analyses

#### eTable 7: Neuroimaging model outcomes after adjusting for total intracranial volume as an additional covariate.

| Genetic Group | ROI | Model Outcomes |
|---|---|---|
| *C9orf72* | Total brain | Group: *b*=-25305.1, SE=14635.9, *p*=0.095<br>TIV: *b*=0.2, SE=0.02, *p*<0.0001<br>Group-by-age: *b*=-1651.8, SE=1766.2, *p*=0.35 |
| | Frontal pole | Group: *b*=-6.92, SE=57.8, *p*=0.91<br>TIV: *b*=0.0004, SE=0.00019, *p*=0.05<br>Group-by-age: *b*=1.75, SE=16.4, *p*=0.92 |
| | Medial orbitofrontal cortex | Group: *b*=-318.1, SE=184.1, *p*=0.095<br>TIV: *b*=0.003, SE=0.0006, *p*<0.0001<br>Group-by-age: *b*=77.4, SE=54.5, *p*=0.16 |
| | Thalamus | Group: *b*=-788.5, SE=259.8, ***p*=0.0054<br>TIV: *b*=0.005, SE=0.0008, *p*<0.0001<br>Group-by-age: *b*=-13.1, SE=68.3, *p*=0.85 |
| | Insula | Group: *b*=-231.0, SE=225.2, *p*=0.31<br>TIV: *b*=0.003, SE=0.0006, *p*<0.0001<br>Group-by-age: *b*=-97.9, SE=52. 9, *p*=0.07 |
| | Rostral anterior cingulate cortex | Group: *b*=-216.5, SE=170.1, *p*=0.21<br>TIV: *b*=0.0017, SE=0.0004, *p*=0.0001<br>Group-by-age: *b*=52.6, SE=30.4, *p*=0.088 |
| GRN | Total brain | Group: *b*=-6109.2, SE=11620.5, *p*=0.61<br>TIV: *b*=0.65, SE=0.05, *p*<0.0001<br>Group-by-age: *b*=2555.4, SE=2696.4, *p*=0.35 |
| | Frontal pole | Group: *b*=25.6, SE=59.3, *p*=0.67<br>TIV: *b*=0.0005, SE=0.0002, *p*=0.05<br>Group-by-age: *b*=11.3, SE=15.2, *p*=0.46 |
| | Medial orbitofrontal cortex | Group: *b*=284.0, SE=268.4, *p*=0.31<br>TIV: *b*=0.004, SE=0.0009, *p*<0.0001<br>Group-by-age: *b*=8.68, SE=51.0, *p*=0.87 |
| | Insula | Group: *b*=11.3, SE=287.7, *p*=0.97<br>TIV: *b*=0.004, SE=0.0012, *p*=0.0028<br>Group-by-age: *b*=13.0, SE=55.6, *p*=0.82 |
| | Rostral anterior cingulate cortex | Group: *b*=173.9, SE=147.6, *p*=0.26<br>TIV: *b*=0.003, SE=0.0005, *p*<0.0001<br>Group-by-age: *b*=25.4, SE=28.7, *p*=0.38 |
| | Putamen | Group: *b*=-50.4, SE=222.7, *p*=0.82<br>TIV: *b*=0.002, SE=0.0008, *p*=0.08<br>Group-by-age: *b*=-23.6, SE=41.8, *p*=0.58 |
| *MAPT* | Total brain | Group: *b*=1512.3, SE=13431.1, *p*=0.91<br>TIV: *b*=0.4, SE=0.04, *p*<0.0001<br>Group-by-age: *b*=-1961.0, SE=1294.4, *p*=0.13 |
| | Frontal pole | Group: *b*=-0.02, SE=70.5, *p*=1.00<br>TIV: *b*=0.0003, SE=0.0002, *p*=0.27<br>Group-by-age: *b*=1.91, SE=10.7, *p*=0.86 |
| | Medial orbitofrontal cortex | Group: *b*=34.9, SE=192.1, *p*=0.86<br>TIV: *b*=0.005, SE=0.0006, *p*<0.0001<br>Group-by-age: *b*=-37.4, SE=28.1, *p*=0.19 |
| | Anterior Medial Temporal Lobe | Group: *b*=-26.790, SE=410.4509, *p*=0.9485<br>TIV: *b*=0.008, SE=0.0014, *p*<0.0001<br>Group-by-age: *b*=-42.856, SE=57.5563, *p*=0.4582 |

**eTable 8: Neuroimaging model outcomes after adjusting for *TMEM106b-rs1990622* genotype as an additional covariate.**

| Genetic Group | ROI | Model Outcomes |
|---|---|---|
| *C9orf72* | Total brain | Group: *b*=-41951.2, SE=26836.8, *p*=0.14<br>*TMEM106B-rs1990622*: *b*=-3721.1, SE=20518.5, *p*=0.86<br>Group-by-age: *b*=2314.4, SE=2752.5, *p*=0.40 |
| | Frontal pole | Group: *b*=-23.2, SE=72.6, *p*=0.75<br>*TMEM106B-rs1990622*: *b*=7.54, SE=53.9, *p*=0.89<br>Group-by-age: *b*=7.10, SE=19.9, *p*=0.72 |
| | Medial orbitofrontal cortex | Group: *b*=-189.7, SE=293.5, *p*=0.53<br>*TMEM106B-rs1990622*: *b*=-79.9, SE=218.1, *p*=0.72<br>Group-by-age: *b*=142.3, SE=71.9, *p*=0.053 |
| | Thalamus | Group: *b*=-1246.7, SE=442.4, **p*=0.011<br>*TMEM106B-rs1990622*: *b*=52.8, SE=334.5, *p*=0.88<br>Group-by-age: *b*=165.5, SE=103.1, *p*=0.11 |
| | Insula | Group: *b*=-799.8, SE=362.8, **p*=0.041<br>*TMEM106B-rs1990622*: *b*=182.2, SE=272.9, *p*=0.51<br>Group-by-age: *b*=37.4, SE=73.7, *p*=0.61 |
| | Rostral anterior cingulate cortex | Group: *b*=-394.5, SE=247.4, *p*=0.13<br>*TMEM106B-rs1990622*: *b*=46.3, SE=187.1, *p*=0.81<br>Group-by-age: *b*=79.2, SE=38.9, **p*=0.047 |
| GRN | Total brain | Group: *b*=27791.0, SE=37455.0, *p*=0.48<br>*TMEM106B-rs1990622*: *b*=41963.3, SE=31530.1, *p*=0.22<br>Group-by-age: *b*=-672.9, SE=2128.1, *p*=0.75 |
| | Frontal pole | Group: *b*=-6.27, SE=60.7, *p*=0.92<br>*TMEM106B-rs1990622*: *b*=170.8, SE=67.1, **p*=0.03<br>Group-by-age: *b*=16.2, SE=17.2, *p*=0.35 |
| | Medial orbitofrontal cortex | Group: *b*=375.6, SE=390.2, *p*=0.36<br>*TMEM106B-rs1990622*: *b*=392.8, SE=342. 2, *p*=0.28<br>Group-by-age: *b*=-7.16, SE=56.2, *p*=0.90 |
| | Insula | Group: *b*=172.6, SE=285.1, *p*=0.56<br>*TMEM106B-rs1990622*: *b*=300.1, SE=334.8, *p*=0.40<br>Group-by-age: *b*=-23.1, SE=53.5, *p*=0.67 |
| | Rostral anterior cingulate cortex | Group: *b*=324.3, SE=232.7, *p*=0.20<br>*TMEM106B-rs1990622*: *b*=500.1, SE=213.6, **p*=0.047<br>Group-by-age: *b*=5.48, SE=27.5, *p*=0.84 |
| | Putamen | Group: *b*=166.6, SE=262.9, *p*=0.54<br>*TMEM106B-rs1990622*: *b*=334.3, SE=276.2, *p*=0.26<br>Group-by-age: *b*=-40.9, SE=47.2, *p*=0.39 |
| *MAPT* | Total brain | Group: *b*=-36242.8, SE=25832.9, *p*=0.18<br>*TMEM106B-rs1990622*: *b*=-15042.8, SE=18639.5, *p*=0.43<br>Group-by-age: *b*=-3225.4, SE=1250.4, *p*=0.012 |
| | Frontal pole | Group: *b*=-90.2, SE=76.5, *p*=0.25<br>*TMEM106B-rs1990622*: *b*=34.2, SE=55.0, *p*=0.54<br>Group-by-age: *b*=-5.79, SE=11.2, *p*=0.61 |
| | Medial orbitofrontal cortex | Group: *b*=-358.5, SE=301.8, *p*=0.25<br>*TMEM106B-rs1990622*: *b*=22.0, SE=217.1, *p*=0.92<br>Group-by-age: *b*=-57.3, SE=32.7, *p*=0.08 |
| | Anterior Medial Temporal Lobe | Group: *b*=-356.4, SE=517.9, *p*=0.50<br>*TMEM106B-rs1990622*: *b*=350.7, SE=380.7, *p*=0.37<br>Group-by-age: *b*=-56.3, SE=61.9, *p*=0.37 |

**eTable 9: Executive function composite score model outcomes after adjusting for site as an additional covariate.**

| Genetic Group | Model Outcomes |
|---|---|
| *C9orf72* | Group: *b*=0.08, SE=0.14, *p*=0.57<br>Group-by-age: *b*=-0.04, SE=0.03, *p*=0.26 |
| GRN | Group: *b*=-0.05, SE=0.17, *p*=0.76<br>Group-by-age: *b*=0.004, SE=0.04, *p*=0.92 |
| *MAPT* | Group: *b*=0.10, SE=0.20, *p*=0.63<br>Group-by-age: *b*=-0.007, SE=0.03, *p*=0.82 |
